# Integrated Clinicogenomic Risk Modeling for Metachronous Second Primary Cancers

**DOI:** 10.64898/2026.06.12.26355388

**Authors:** Johnathan Amsalem, Irina Ostrovnaya, Andrew R. Marderstein, Ying L. Liu, Tomin Perea-Chamblee, Vignesh Ravichandran, Justin Jee, Michael Conry, Aliya Khurram, Yelena Kemel, Ellen Kim, Semanti Mukherjee, Alicia Latham, Lauren Banaszak, Ritika Kundra, Saibaba Magunta, Christopher Fong, Matthew F. Buas, Chaitanya Bandlamudi, Jonine Bernstein, Venkatraman Seshan, Gilles Salles, Diana Mandelker, Michael F. Berger, David B. Solit, Zsofia K. Stadler, Jian Carrot-Zhang, Nikolaus Schultz, Kenneth Offit, Vijai Joseph

## Abstract

Improvements in cancer survival have increased the burden of subsequent primary malignancies. We developed and validated a programmatic classifier of multiple primary cancers (MPC) to derive second cancer phenotypes at scale. Among 81,175 cancer patients, we identified 56 first-second cancer pairs, 22 of which exceeded SEER primary cancer incidence rates. Even after accounting for various known risk factors, substantial elevated risk persisted, even in established hereditary cancer pairs (breast-ovary, breast-pancreas, prostate-pancreas), suggesting that current screening protocols do not adequately account for MPC susceptibility. To address this limitation, we built machine-learning models integrating rare germline variants, polygenic risk scores, treatment exposures, and demographic features to predict site-specific second primaries in breast and prostate cancer survivors. These models accurately predicted second ovarian and pancreatic cancers across a long follow-up period (15-year time-dependent AUC 0.70). This is the first systematic, pan-cancer integration of clinicogenomic factors for early prediction of second-primary malignancies. Our framework enables individualized risk estimation, enhanced targeted surveillance, and cancer prevention amongst a growing population of cancer survivors.

**Statement of Significance:** We identified second cancers that occurred more often than expected among survivors. Predictive models using genetic, lifestyle, and clinical factors accurately identified patients at higher risk of second hereditary cancers. Such predictions can enable cost-effective, selective surveillance in a growing population of cancer survivors, reducing cancer burden.

## Introduction

Five-year survival across all cancers has risen to 70%, a significant increase from 50% in the mid-1970s [1]. Improvements in cancer detection and treatment have led to a growing population of cancer survivors. However, approximately one in five individuals will develop a second primary cancer over their lifetime, a burden expected to increase as outcomes continue to improve [2–4]. For survivorship care, the unmet need is not only to describe this burden, but to identify which individuals are at highest risk, particularly for second cancers which may be due to hereditary predisposition, treatment, or specific exposures, where earlier detection of an anticipated second cancer may be consequential.

A central challenge to both etiologic studies and risk stratification of second cancers is defining the outcome itself, i.e., distinguishing true second primary malignancies from recurrences and metastases [5–6]. While it is often possible to compare primary and second cancers using histology, medical records, and imaging, large-scale classification remains challenging in real-world cohorts. Care across institutions, variable longitudinal documentation, and site- or histology-specific nomenclature ambiguities can introduce systematic misclassification of multiple primary cancers (MPC). Such misclassification can introduce bias into population-scale estimates of MPC risk and limit the development of reliable predictive models, although recent integrative approaches combining clinical and molecular data have begun to define these limitations [7,8].

To address this gap, we first developed a programmatic approach that implements International Agency for Research on Cancer (IARC)-informed classification of cancer diagnoses as primary or non-primary using structured tumor registry variables together with relevant clinical context. This approach is designed to approximate expert clinical reasoning by integrating standardized registry variables and clinical data. By leveraging structured data elements, our method enables scalable, automated classification of MPC without requiring manual chart review.

Using this infrastructure, we next conducted a population-scale analysis of MPC to characterize patterns of excess second cancer risk. We quantified risk across cancer-pair combinations using standardized incidence ratios (SIR) and then assessed heterogeneity by integrating germline pathogenic variants (PV), imputed polygenic risk scores (PRS), treatment exposures, and clinical covariates. Clinical covariates, including age at first cancer diagnosis, smoking status, body mass index (BMI), and treatment exposures, were derived from the Memorial Sloan Kettering (MSK) Cancer Data Science Initiative, which harmonizes electronic medical records (EMR), revenue management system data, and tumor registry records using natural language processing (NLP) and regular expression extraction [8].

Although many of these analyses recapitulated established etiologic contributors to second-cancer risk, substantial residual risk remained across numerous cancer pairs, including syndromic and hereditary tumor types, suggesting a multifactorial basis for MPC susceptibility. This heterogeneous risk architecture motivated complementing inferential analyses with predictive modeling aimed at survivor-level risk stratification rather than a single-factor explanation. We therefore developed a model referred to herein as *Predictive Risk Estimation to Detect Independent Cancer Types* (“MPC-PREDICT”), a set of penalized Fine-Gray subdistribution hazard models using Least Absolute Shrinkage and Selection Operator (LASSO) regularization [9,10] to integrate clinical, exposure, germline, and treatment features. This approach directly models risk in the presence of competing events, such as death, while enabling time-to-event prediction and feature selection in high-dimensional settings. To our knowledge, no prior framework has integrated this breadth of clinicogenomic data types to predict specific second primary cancers in a clinical cohort of this scale. By combining scalable MPC classification with risk estimation in cancer survivors, we demonstrate that second primary cancer susceptibility can be estimated for individual patients, even in the absence of a single dominant factor, supporting predictive, risk-adapted surveillance strategies in an expanding survivor population.

## Materials and Methods

### Cohort Selection

81,175 consented patients from the MSK clinical sequencing cohort (MSK-IMPACT), all of whom underwent tumor-normal (T:N) sequencing [11], were selected. Cancer records from these patients were processed by the programmatic classifier. Of these, 821 patients were excluded due to missing tumor diagnosis data (topography or morphology codes) or because none of their recorded neoplasms met criteria for a primary malignancy. Synchronous cancer pairs (defined as cancers diagnosed within 1 year) were also excluded to prevent potential surveillance-bias, resulting in a final analytical cohort of 77,060 patients. All analyses were conducted in accordance with institutional ethical standards and were approved by the MSK Institutional Review Board (protocols 16-1249 and 18-420).

### MPC Classifier Design

We developed a programmatic, rule-based classifier to identify MPC by distinguishing recurrences, metastatic, and non-primary/non-malignant neoplasms from primary tumors using structured tumor registry data. Tumors were standardized using ICD-O-3 topology and morphology codes [12] and then processed by the classifier. Records classified as non-primary (e.g., *in situ* neoplasms or metastasis) were excluded from the classifier’s output but retained for performance evaluation. The remaining tumors were then selected, assigning each patient a primary cancer count, and classifying them as having a single primary (SP) or multiple primary cancers (MPC).

Classification was performed at the patient level, incorporating the temporal ordering of tumor records, and was grounded in IARC’s multiple primary cancer rules [13], with clinically informed extensions to address common registry ambiguities encountered in real-world oncology data [14]. These included handling of paired organs, related anatomic subsites, and systemic malignancies, as well as temporal separation criteria to distinguish late second primaries from recurrence. The framework was designed to be conservative, collapsing tumors into a single primary when evidence for independence was insufficient, thereby avoiding inflation of MPC counts.

The classifier was implemented using Python 3.13. Detailed rule logic and exception handling are provided in the **<u>Supplementary Materials and Methods</u>**.

### Classifier Validation of MPC Status

The MPC classifier’s performance was evaluated using a pre-curated subset of the inception cohort (N = 81,175) comprising >25,000 consented Memorial Sloan Kettering Cancer Center (MSKCC) patients with longitudinal tumor histories. Every patient in this subset had at least one primary cancer that underwent expert-adjudication by two or more clinicians through manual chart review [15]. This dataset served as a “*truth set*.” All patients were included in performance evaluation, including those labeled as having no primary cancers by the programmatic classifier (e.g., patients with *in situ* tumors only).

Classifier outputs were compared with expert adjudication at two levels as described in **Supplementary Materials and Methods.**

Following validation of the classifier’s performance, we applied it to the inception cohort (N = 81,175), all of which had available tumor registry data. All downstream analyses excluded synchronous cancer pairs, as concurrent detection may reflect surveillance or detection bias.

### Cumulative Incidence Analysis

Using MPC classifications derived from the automated classifier, cumulative incidence functions for metachronous second primary cancers (>1-year after first cancer) and death were estimated using nonparametric competing-risk methods [16]. Time-to-event follow-up began one year after the first primary cancer diagnosis (1-year landmark), and individuals were required to be event-free at this time. Follow-up continued until a metachronous second primary malignancy, death, or censoring at last clinical follow-up. Same-site cancer pairs were excluded to minimize misclassification. Cumulative incidence estimates, accounting for competing mortality, were summarized at prespecified time horizons of 5, 10, and 15 years, stratified by common first primary cancer types.

### Standardized Incidence Ratios (SIR)

To complement absolute burden estimates with site-specific risk, standardized incidence ratios (SIR) were estimated for 56 cancer-pair combinations using age- and sex-specific cancer incidence rates from the SEER-21 registry [17], following a previously described approach [15], as described in **<u>Supplementary Materials and Methods.</u>** Sensitivity analyses were performed by re-estimating SIR without latency weighting to assess the robustness of excess risk estimates <u>(**Supplementary Tables 1 and 2**)</u>.

### Risk Stratification by Clinical and Genetic Factors

Metachronous cancer pairs with excess risk identified in the primary SIR analysis were further evaluated by stratifying the cohort according to clinical, genetic, and treatment-related factors. Within each stratum, SIRs were estimated to assess heterogeneity. In analyses requiring adjustment for multiple covariates or incorporation of time-dependent effects, regression models were additionally employed.

Smoking status (never vs. ever) and body mass index (BMI; normal, overweight, obese) were examined using stratum-specific SIRs with pre-specified comparisons across categories.

Germline genetic risk was assessed in selected hereditary cancer pairs (breast-ovary, breast-pancreas, and prostate-pancreas) using pathogenic or likely pathogenic (PLP) variants in cancer susceptibility genes, with patients classified by gene penetrance as high risk, moderate/low risk, or non-carriers. In parallel, polygenic risk scores (PRS) were used to evaluate common genetic predisposition, comparing second cancer risk across PRS deciles using Fine-Gray regression.

Age at first cancer diagnosis was analyzed both categorically (quintiles) and continuously (using Fine-Gray regression) to estimate associations with second primary cancer risk. Additional case-only logistic regression analyses were performed to assess robustness of risk estimates, and linear regression was used to evaluate associations with latency to second cancer.

Treatment exposures were modeled as time-dependent covariates to evaluate associations between selected therapies and second primary cancers, with adjustment for demographic, clinical, and treatment-related factors.

Additional methodological details, including genetic variant definitions, PRS construction, sensitivity analyses, and model specifications, are provided in the **<u>Supplementary Materials and Methods</u>**.

### LASSO Fine-Gray Regression for Prediction of Hereditary Cancer Pairs

MPC-PREDICT modeling focused on three hereditary cancer pairs (breast-ovary, breast-pancreas, and prostate-pancreas) in patients with germline consent and available genotype data (n = 32,200; **<u>Figure 1</u>**). This selection was driven by the high frequency of metachronous second cancers in breast and prostate cancer survivors (**<u>Supplementary Figure 1</u>**), relatively favorable survival outcomes that allowed sufficient time at risk (**<u>Supplementary Table 3</u>**), elevated excess risk (**<u>Figure 3</u>**), and enough events among germline-consented patients to support multivariable modeling.

**Figure 1.**
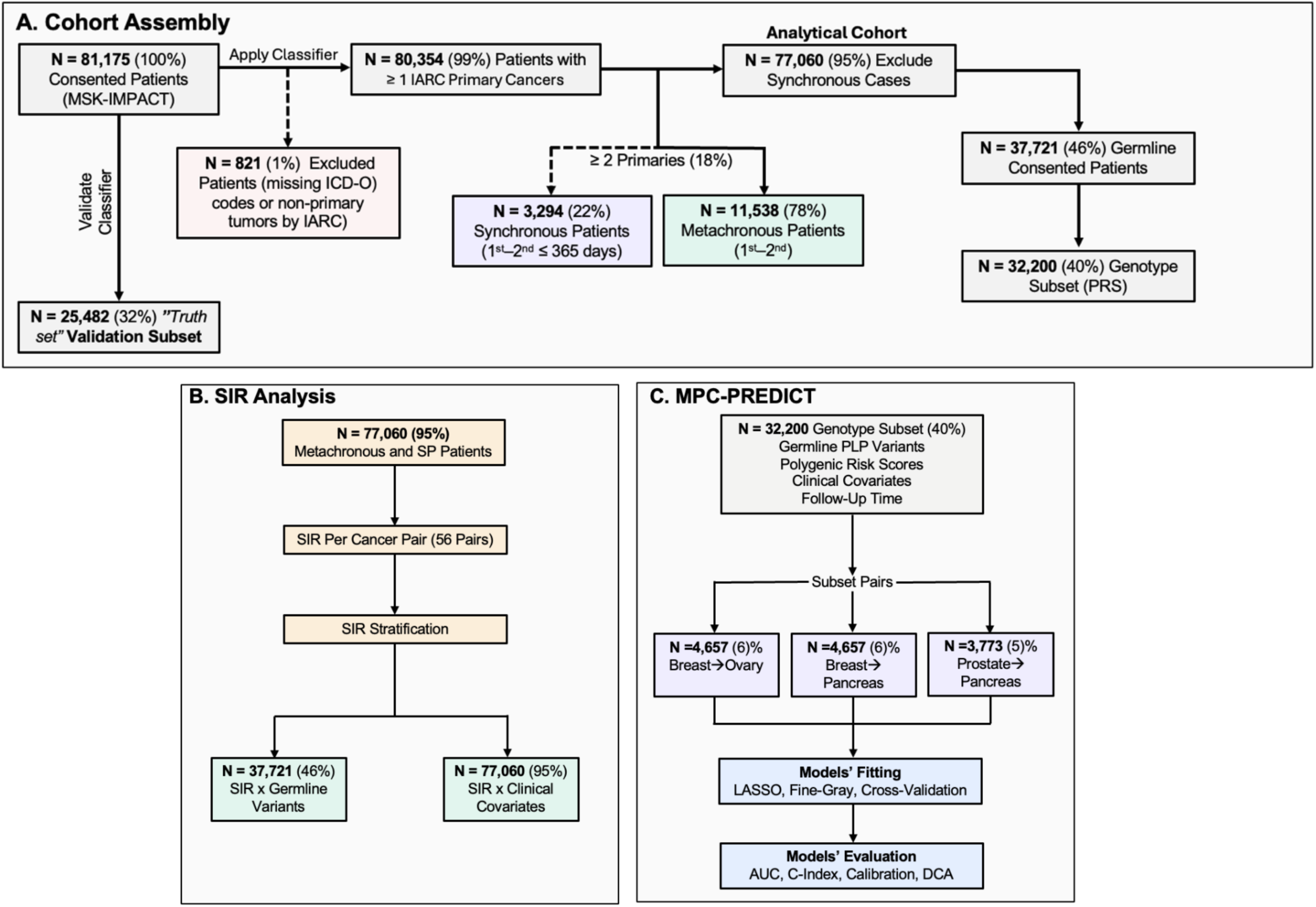
Study cohort and design. (A) Cohort Assembly. Flow diagram of the inception cohort of 81,175 consented patients in the MSK-IMPACT study. After excluding 821 patients with missing ICD-O codes or non-primary tumors (per IARC criteria), 80,354 patients with ≥1 primary cancers were included in the cohort. Of these, 3,294 had synchronous multiple primary cancers (diagnosed within one year of the first cancer) and were excluded from downstream analyses due to potential surveillance bias (e.g. SIR). The remaining 77,060 patients comprised the analytical cohort, including 11,538 with metachronous second primaries (diagnosed more than one year after the first) and 65,522 with a single primary cancer. Among these, 37,721 had consented to receive results of germline testing, of whom 32,200 also had available genotype data for polygenic risk score analysis and risk modeling. Additionally, a subset of 25,482 patients formed a validation subset used to assess classifier’s performance. (B) SIR Analysis. SIR analyses were conducted across 56 cancer pairs among the analytical cohort of 77,060 patients. Follow-up analyses stratified SIR by clinical covariates (e.g., smoking, treatment, age) and germline PLP variant status (among the 37,721 germline-consented patients), to identify factors driving excess second primary cancer risk. (C) MPC-PREDICT. Modeling was performed in the genotype subset of 32,200 patients (40%), incorporating germline PLP variants, polygenic risk scores, clinical covariates, and follow-up time. Analyses were restricted to three hereditary cancer pairs: breast-to-ovary (n = 4,657; 6%), breast-to-pancreas (n = 4,657; 6%), and prostate-to-pancreas (n = 3,773; 5%). Models were fit for each cancer pair individually using LASSO and Fine-Gray regression methods with cross-validation, and evaluated by AUC, C-index, calibration, and decision curve analysis. *All percentages (%) shown reflect the proportion of the full inception cohort (N = 81,175). Solid arrows denote subset cohorts; dashed arrows denote excluded subgroups. Abbreviation: PLP, pathogenic or likely pathogenic*.

Separate penalized Fine-Gray regression models using LASSO (MPC-PREDICT) were fit for each predefined hereditary cancer pair. LASSO regularization provided a simple machine-learning framework for joint feature selection and mitigation of overfitting in the presence of correlated clinical, genetic, and treatment-related covariates [9].

For each cancer pair, the primary endpoint was defined as the incidence of the second cancer of interest (ovary/pancreas). Follow up started 1-year after the first primary cancer diagnosis (1-year landmark), to exclude synchronous pairs, and continued until the development of the second metachronous cancer of interest, a competing event (including death or non-target second primary cancers), or censoring at last clinical follow-up. Fine–Gray regression models were used to estimate the absolute risk of second primary cancers, accounting for competing risks such as death or other second cancers. All predictors were defined using information available up to the 1-year landmark to prevent information leakage, with baseline variables (e.g., germline genetic factors) included as fixed covariates.

MPC-PREDICT models were trained using a 75%, 25% training–validation split. Within the training set, Monte Carlo 3:1 cross-validation (100 iterations) [18] was performed using repeated random subsampling, in which penalized Fine-Gray models were fit across a grid of λ values and evaluated on held-out subsets of the training data. Model performance was evaluated on the validation split after finalizing λ from training. Discrimination, calibration and clinical relevance were evaluated as described in **<u>Supplementary Materials and Methods.</u>**

### Statistical Software

All statistical analyses were performed using R version 4.5.1. PRS were generated using PRSice- 2 [19]. MPC-PREDICT penalized Fine-Gray regression with LASSO regularization was implemented using the *fastcmprsk* package in R [20]. Time-dependent AUC and calibration were assessed using the *riskRegression* package [21], and clinical benefit was tested using the *dcurves* package in R [22].

### Data Availability

Portions of the data underlying this study are available through controlled or public repositories [23]. Individual-level data from the full cohort are not publicly available due to patient privacy considerations. Summary-level and derived data supporting the findings are provided in the Supplementary Data and/or are available from the corresponding author upon reasonable request. Code used for analysis will be made available on GitHub.

## Results

### Cohort Selection

An inception cohort of 81,175 patients consented to MSK-IMPACT testing, 821 of which were excluded by the MPC classifier (**<u>Figure 1A</u>**), leaving 80,354 patients. Among these, 3,294 had synchronous cancer pairs and were further excluded, yielding a final analytical cohort of 77,060 patients. The analytical cohort was 53% female, with a median age at first cancer of 59 years. Across this analytical cohort, 91,267 primary tumors were identified, with breast (12%), lung (11%), and prostate (8%) cancers being the most common.

Overall, 15% (n = 11,538) of the analytical cohort had metachronous MPC. Of these, 81% had two primary cancers, 15% had three, and 4% had four or more. The distributions of these cancer pairs are shown in **<u>Supplementary Figure 1</u>**. The median ages at first and second cancer diagnoses among MPC patients were 58 and 68 years, respectively. First cancers in metachronous MPC patients were diagnosed at earlier stages than SP (23.9% vs 10.7% stage 0-II, **<u>Table 1</u>**), consistent with their improved survival, which allows time for second cancers to develop.

**Table 1.** Patient characteristics stratified by multiple primary cancer status and germline testing results. Patient characteristics stratified by multiple primary cancer status and germline testing results among the full analytical cohort of 77,060 patients. Values are presented as n (%) for categorical variables and as median (interquartile range) for continuous variables. The comparison between PV carriers and non-carriers is restricted to the subset of patients who underwent germline testing and excludes synchronous cancer pairs (n = 37,721); non-carriers tested negative for all pathogenic variants on the panel. Ancestry, ethnicity, stage, and BMI had missing values, and participants missing these variables were excluded from the corresponding analyses. P-values compare single primary (SP) versus multiple primary cancer (MPC) patients and, separately, PV carriers versus PV-negative patients, using Fisher’s exact tests for categorical variables and Wilcoxon rank-sum tests for continuous variables. *Abbreviation: PV, pathogenic or likely pathogenic variant; SP, single primary; MPC, multiple primary cancers*.

|  | SP (N = 65,522) | Metachronous MPC (N =11,538) | P-Value | PV Carriers (N=6,324) | Non-Carriers (N=31,397) | P-value |
| --- | --- | --- | --- | --- | --- | --- |
| **Sex (%)** |  |  |  |  |  |  |
| Female | 34,939 (53.3%) | 6,187 (53.6%) | 0.557 | 3,485 (55.1%) | 17,404 (55.4%) | 0.134 |
| Male | 30,583 (46.7%) | 5,351 (46.4%) |  | 2,839 (44.9%) | 13,993 (44.6%) |  |
| **Ethnicity (%)** |  |  |  |  |  |  |
| Hispanic | 5,106 (8.2%) | 565 (5.2%) | < 0.001 | 466 (7.4%) | 2,717 (9.1%) | <0.001 |
| Non-Hispanic | 57,060 (91.8%) | 10,369 (94.8%) |  | 5,526 (92.6%) | 26,888 (90.9%) |  |
| **Median BMI (kg/m<sup>2</sup>, IQR)** |  |  |  |  |  |  |
| Female | 25.8 (22.3-30.5) | 26.4 (22.9-31.1) | < 0.001 | 25.8 (22.1-30.5) | 26.2 (22.5-31.1) | < 0.001 |
| Male | 27.3 (24.3-30.7) | 27.8 (25.0-31.1) | < 0.001 | 27.2 (24.2-30.4) | 27.3 (24.3-30.9) | 0.056 |
| **Age At First Diagnosis (Years, IQR)** |  |  |  |  |  |  |
|  | 59.4 (47.8-68.3) | 57.7 (47.5-65.8) | < 0.001 | 56.6 (44.1-65.9) | 59.1 (47.5-67.9) | < 0.001 |
| **Smoker (%)** |  |  |  |  |  |  |
| Current/Former | 22,626 (34.5%) | 5,032 (43.6%) | < 0.001 | 2,139 (33.8%) | 10,557 (33.6%) | 0.042 |
| Never | 33,979 (51.9%) | 5,390 (46.7%) |  | 3,842 (60.8%) | 17,851 (56.9%) |  |
| Unknown | 8,917 (13.6%) | 1,116 (9.7%) |  | 343 (5.4%) | 2,989 (9.5%) |  |
| **Ashkenazi Jewish** |  |  |  |  |  |  |
| Yes | 9,247 (14.5%) | 2,416 (21.3%) | < 0.001 | 1,450 (23.0%) | 4,342 (13.8%) | < 0.001 |
| No | 54,617 (85.5%) | 8,914 (78.7%) |  | 4,866 (77.0%) | 27,027 (86.2%) |  |
| **European** |  |  |  |  |  |  |
| Yes | 37,776 (59.2%) | 7,050 (62.2%) | < 0.001 | 3,592 (56.9%) | 18,344 (58.5%) | 0.019 |
| No | 26,088 (40.8%) | 4,280 (37.8%) |  | 2,724 (43.1%) | 13,025 (41.5%) |  |
| **Stage of First Cancer** |  |  |  |  |  |  |
| Early (0-II) | 6,432 (10.7%) | 1,315 (23.9%) | < 0.001 | 660 (12.4%) | 3,252 (11.9%) | 0.321 |
| Late (III-IV) | 53,708 (89.3%) | 4,180 (76.1%) |  | 4,678 (87.6%) | 24,127 (88.1%) |  |
| **Metachronous MPC (≥2)** |  |  |  |  |  |  |
| Yes | - | - | - | 1,159 (18.3%) | 4,195 (13.4%) | < 0.001 |
| No | - | - | - | 5,165 (81.7%) | 27,202 (86.6%) |  |

Of the analytical cohort, 49% (n = 37,721) consented to receive germline testing results (**<u>Figure 1</u>**). Among those tested, 55% were female (n = 20,889), and 14% had metachronous MPC. Metachronous MPC were more common among carriers than non-carriers (18% vs 13%, Fisher’s exact test p < 0.001; **<u>Table 1</u>**).

We first established the fidelity of MPC phenotyping by validating an IARC-informed classifier against expert adjudication. Using this validated phenotype, we quantified second-cancer excess risk at scale and evaluated the contributions of clinical factors, germline variation, and cancer therapies. Finally, motivated by residual risk unexplained by individual factors, we developed MPC-PREDICT, a set of integrative models to stratify survivor-level risk for metachronous second primaries in hereditary cancer pairs.

### Strong Concordance Between Automated MPC Classification and “Truth Set”

The “*truth set*” comprised of 25,482 consented patients who underwent T:N sequencing and had available manually curated expert-level assertions [15]. The median age at first cancer diagnosis was 57 years (IQR, 45–66). Overall, 55% were female (n = 13,971), 54% were of European ancestry (n = 13,766), and 24% were Ashkenazi Jewish (AJ; n = 6,210). Across this cohort, 30,941 primary tumors were identified, with breast (17%), lung (12%), and colorectal (8%) cancers being the most common.

The classifier showed strong concordance with the *“truth set”,* agreeing on MPC versus SP binary status in 93% of patients (**<u>Figure 2A</u>**). Binary MPC classification showed significant agreement (Cohen’s κ = 0.88; 95% CI, 0.87–0.89), with 81% sensitivity and 100% specificity, consistent with the classifier’s conservative design. Discordant classifications were concentrated in the same cancer types across both binary and tier analyses (**<u>Figure 2B,D</u>**): bladder/urothelial cancers, where *in situ* lesions complicate primary tumor designation; skin cancers, where overlapping histologic subtypes and anatomic proximity can make synchronous tumors difficult to distinguish from recurrence; and breast cancers, where incomplete laterality data frequently obscures tumor independence. Consistent with this, excluding breast cancers improved performance further (accuracy, 98%; κ = 0.90; 95% CI, 0.89–0.91; sensitivity, 84%; **<u>Supplementary Figure 2</u>**).

**Figure 2:**
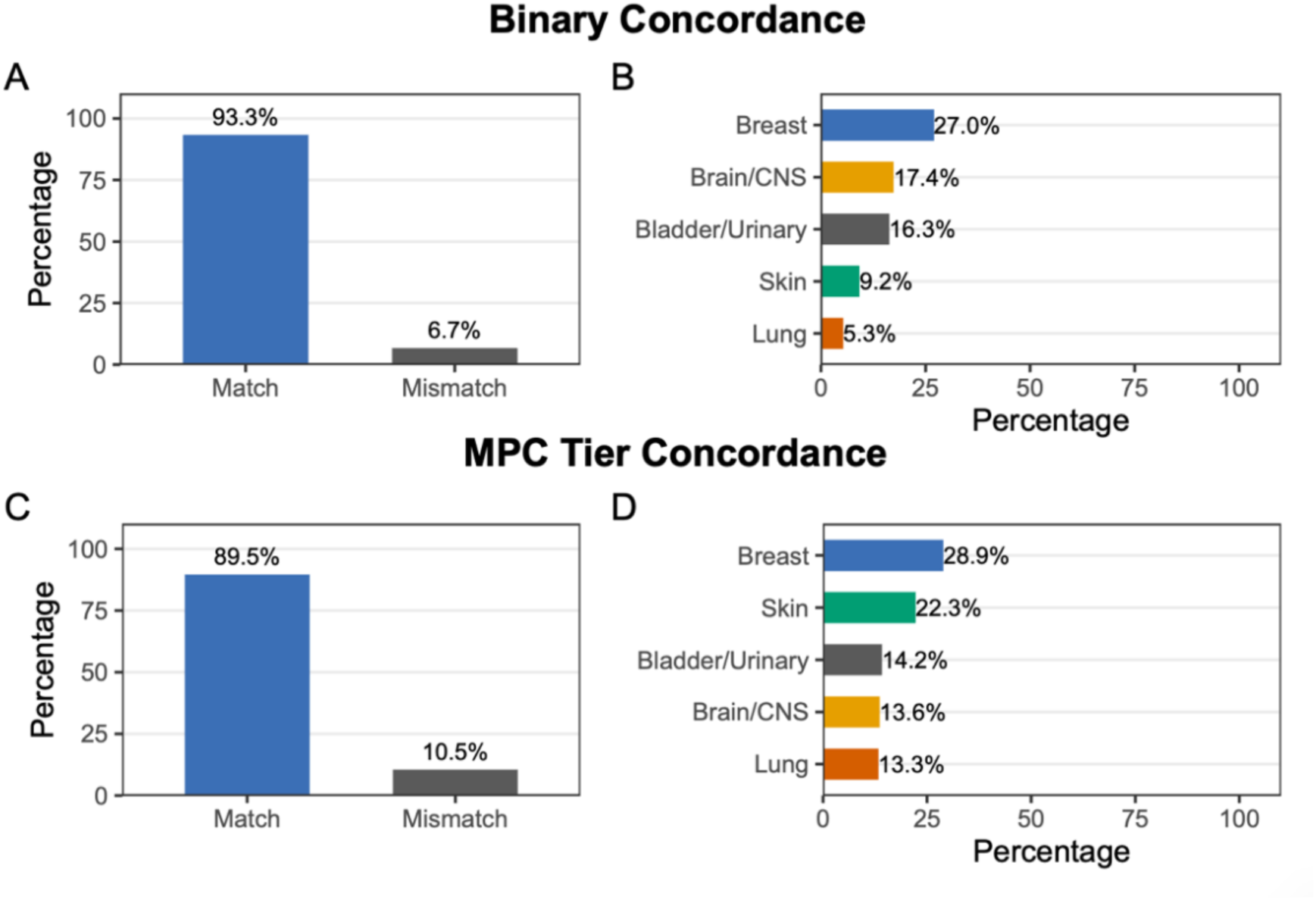
Concordance of classifier assignments with expert-adjudicated “*truth set*” and distribution of mismatch sites. (A) Accuracy for multiple primary cancer (MPC) versus single primary (SP) classification between the classifier and expert review across 25,482 patients. Overall accuracy was 93.3%. (B) Distribution of primary cancer sites among discordant patients with binary classification discrepancies (n = 1,705), highlighting paired organs and sites subject to differences between the algorithm and manual curation. Accuracy of MPC tier assignments (number of distinct primary cancers per patient) across 3,664 MPC patients. Overall accuracy was 89.5% (D) Distribution of cancer sites among tier-discordant cases (n = 383), including paired organs and tumors affected by differences in classification rules.

For MPC tier assignment (the number of primary tumors among patients classified as MPC by both the classifier and expert adjudication), agreement was substantial (weighted κ = 0.78; 95% CI, 0.75–0.80), with an accuracy of 90% (n = 3,281 of 3,664; **<u>Figure 2C</u>**). Performance was stronger for cancer-pair combinations included in the primary SIR analyses, which mainly focus on distinct organ sites and exclude some scenarios where attribution of second primary cancer can be challenging (e.g. breast-breast).

### Age at Diagnosis and Time-Dependent Cumulative Incidence of Metachronous Primary Cancers

We examined age at diagnosis for each cancer type both as a first primary cancer and as a second, metachronous primary cancer within the analytical cohort. Across cancer types, the median age at first primary cancer diagnosis in our cohort was consistently younger than the corresponding median age reported in SEER. Additionally, 6 of the 13 cancer types showed a younger or equivalent age at second cancer diagnosis compared to SEER data **(<u>Supplementary Table 3</u>).**

Accounting for the competing risk of death prior to a second cancer diagnosis, the cumulative incidence of MPC varied markedly by first primary cancer type (**<u>Supplementary Table 3</u>**). Prostate and thyroid cancer survivors had a high absolute burden of subsequent primary malignancies and relatively low mortality prior to a subsequent cancer compared to other tumor types. By 15 years from beginning of follow-up, the cumulative incidence of any second primary cancer reached 38% among prostate cancer survivors and 37% among thyroid cancer survivors. In contrast, ovarian and pancreatic cancer survivors had the lowest 15-year cumulative incidence (11% and 6%, respectively) but higher mortality. Overall, mortality and second cancer rates varied substantially across cancer types (**<u>Supplementary Table 3</u>**) and were generally inversely associated. Poor survival cancers have less follow-up time in which second cancers can occur, while higher survival cancers accrue more person-time at risk. Thus, cumulative incidence reflects a complex interplay of biology and the competing risks, that requires a broader understanding of the multitude of factors at play.

### Wide Spectrum of Second Cancer Excess Risk

To identify specific high-risk cancer pairs, we estimated SIR using age- and sex-specific SEER-21 population-based reference rates across 56 cancer pairs. 22 metachronous cancer pairs with significantly elevated SIR were identified, including 14 in females and 8 in males (**<u>Figure 3</u>**). In females, the largest excess risks included ovary-leukemia (SIR = 6.0), breast-pancreas (SIR = 4.7), melanoma-lung (SIR = 4.3), and breast-ovary (SIR = 3.9). In males, elevated SIR include prostate-pancreas (SIR = 3.7), bladder-lung (SIR = 3.0) and lung-lung (SIR = 2.3). Novel cancer pairs exhibiting excess risk included Non-Hodgkin Lymphoma (NHL)–related sequences, as well as ovarian–leukemia and prostate–leukemia pairs. Sensitivity analyses using unweighted SIR estimates, along with corresponding observed case counts for all cancer pairs are provided in **<u>Supplementary Tables 1 and 2</u>**.

**Figure 3.**
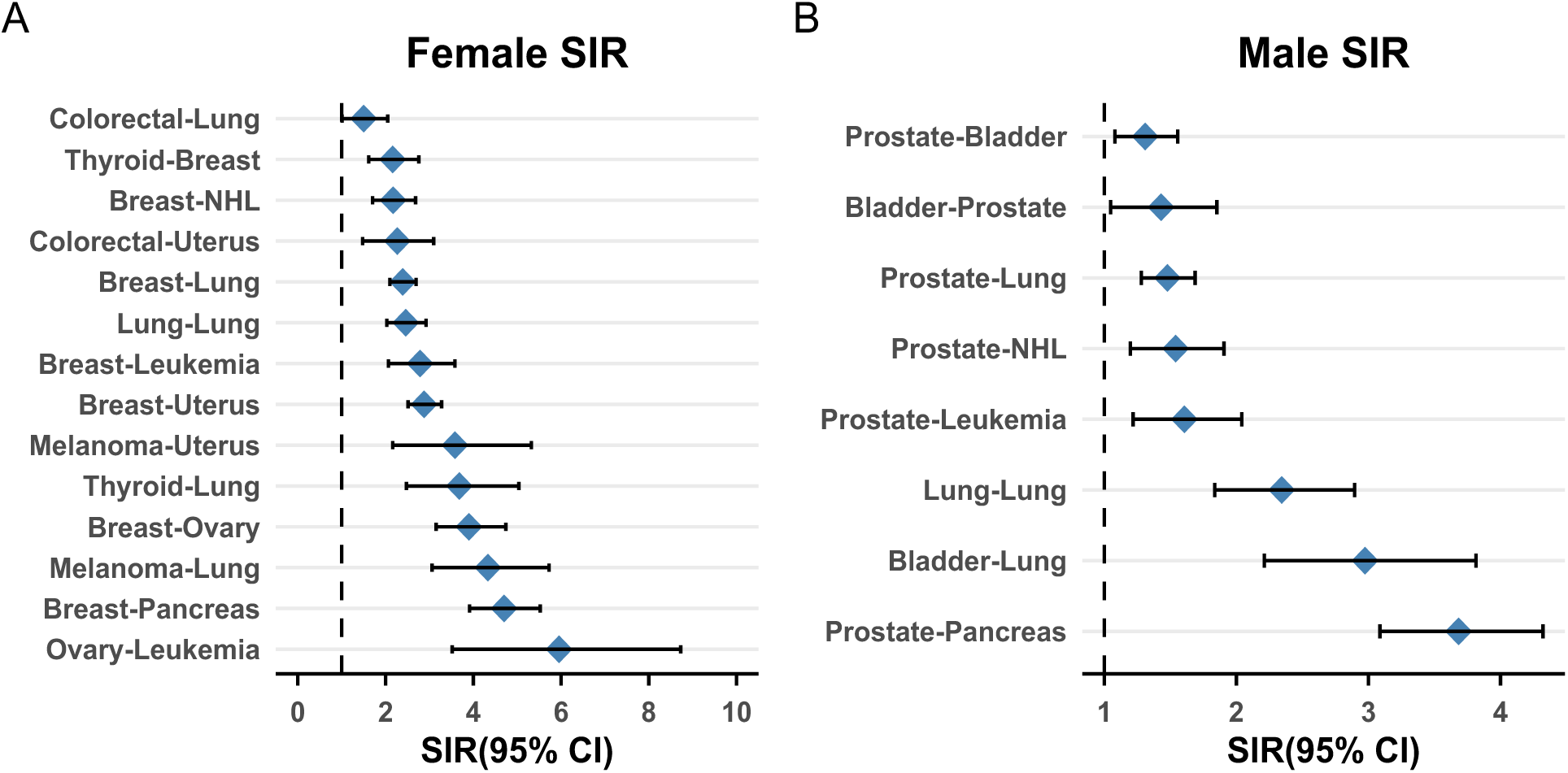
Standardized incidence ratios (SIR) for metachronous cancer-pair combinations in (A) females and (B) males. Each row represents a cancer-pair combination in the format “first cancer - second cancer” ordered by date of diagnosis. The x-axis shows the SIR with 95% bootstrap CI’s, on a linear scale. The vertical dashed line at SIR = 1 denotes the null expectation (no excess or deficit in risk compared to the general population). Points to the right of the line indicate increased risk of the second primary relative to SEER-21 age- and sex-specific incidence rates. A total of 14 cancer-pair combinations were significant in females and 8 in males. *Abbreviations: NHL:Non-Hodgkin Lymphoma*

### Heterogeneous EQects of Clinical, Genetic, and Treatment-Related Risk Modifiers

Stratified analyses, with SIRs calculated relative to the SEER general population, identified multiple clinical, genetic, and treatment-related factors associated with variation in secondary cancer risk across high-risk cancer pairs **(<u>Figure 4</u>**).

**Figure 4.**
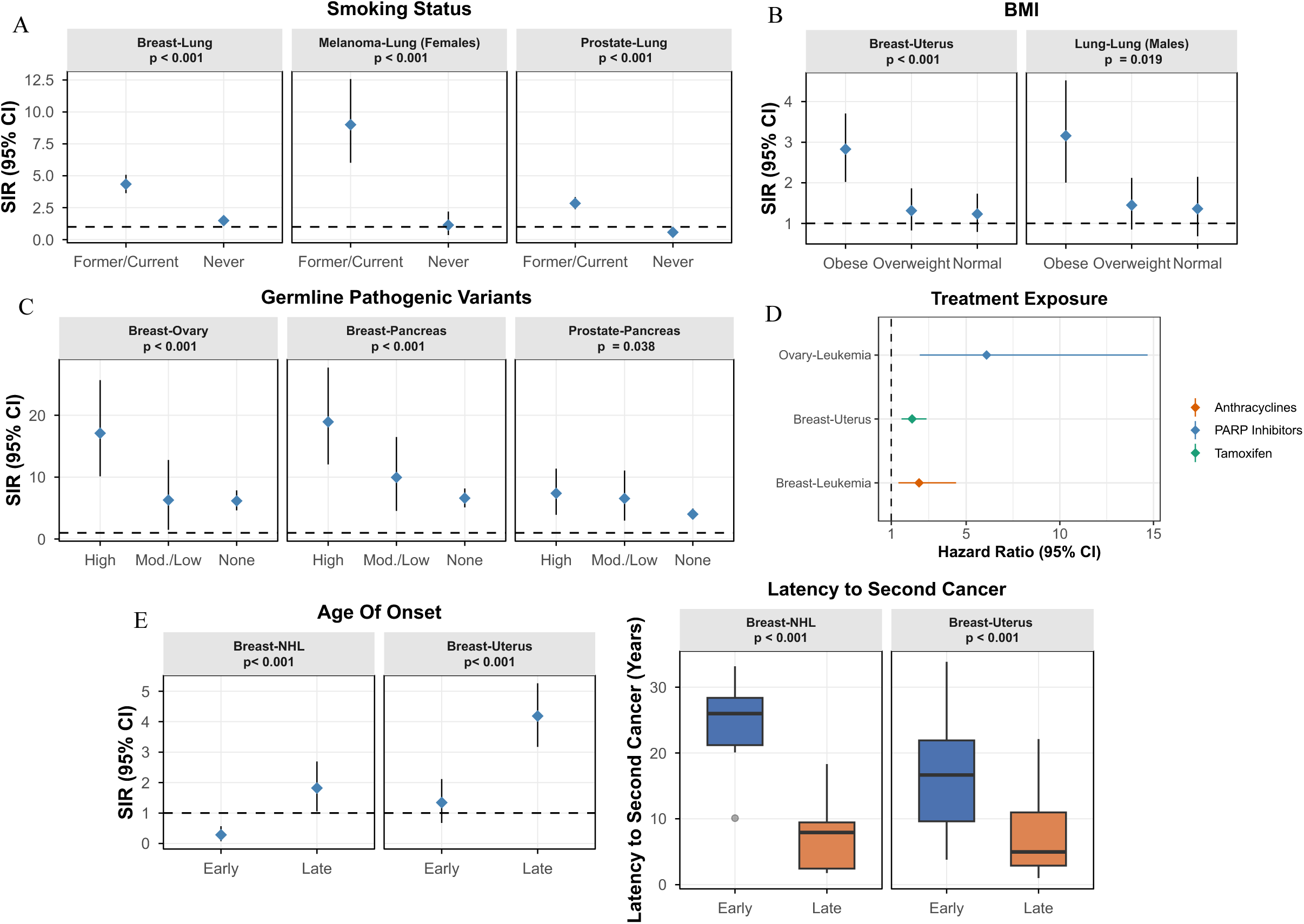
Clinical, genetic, and treatment-related modifiers of secondary cancer risk. (A) Smoking Status: SIR for second primary cancers (Breast-Lung, Melanoma-Lung in females, and Prostate-Lung) stratified by smoking history. Data points represent the SIR with vertical bars indicating the 95% Confidence Interval (CI). The dashed horizontal line represents an SIR of 1.0 (baseline risk). (B) BMI: Comparison of secondary cancer risk (Breast-Uterus and Lung-Lung in males) across BMI categories. (C) Germline pathogenic variants. SIR for canonical hereditary cancer pairs stratified by germline pathogenic variant burden (high, moderate/low, none) across NCCN-defined susceptibility genes. P values were derived from comparing high–penetrance variant carriers with non-carriers. (D) Treatment exposure: Associations between treatment exposures and second cancer risk estimated using time-varying Cox proportional hazards models. (E) Secondary cancer risk and latency stratified by late vs. early (80^th^ percentile vs. 20^th^ percentile) age at first cancer diagnosis for breast-uterus and breast-NHL cancer pairs. Panels display standardized incidence ratios for second primary cancer risk and corresponding latency distributions to second cancer across age-at-onset strata. P-values were derived from Wald tests comparing the log-transformed SIRs. *Abbreviations: NHL:Non-Hodgkin Lymphoma*.

Smoking status strongly modified risk of second lung cancers across various cancer pairs, with former or current smokers exhibiting significantly higher SIR than non-smokers for colorectal-lung, breast-lung, lung-lung, thyroid-lung, and melanoma-lung in females, and prostate-lung, lung-lung, and bladder-lung in males (all p < 0.001).

BMI also stratified risk, with obesity associated with elevated SIR for breast-uterus in females (p < 0.001) and lung-lung cancers in males (p = 0.018).

Older age at first cancer diagnosis (80th vs 20th percentile; late- vs early-onset group) was associated with a higher incidence of second primary cancers for breast-uterus and breast-NHL cancer pairs in females (all p < 0.001). For breast-uterus cancer pairs, the median age at first diagnosis was 39 years in the early-onset group and 69 years in the late-onset group; corresponding values for breast-NHL pairs were 38 and 68 years. Older age at first diagnosis was also associated with shorter latency to second cancer. Median latency was longer in the early-onset group than in the late-onset group for both breast-uterus (17 vs. 5 years) and breast-NHL (26 vs. 8 years) cancer pairs (Wilcoxon rank-sum test, p < 0.001 for both) (**<u>Figure 4E</u>**).

When age at first cancer diagnosis was modeled as a continuous variable, older age was consistently associated with a higher incidence of second primary cancers across multiple cancer pairs in Fine–Gray competing risk models, with all reported associations remaining significant after correction for multiple testing (**<u>Figure 5</u>**). In females, increasing age at first diagnosis was associated with elevated subdistribution hazard ratios (SHRs) for second primary cancers, including breast–pancreas, breast–lung, breast–uterus, breast–NHL, and melanoma–lung, with the strongest effect observed in colorectal–lung, where each standard-deviation increase in age at colorectal cancer diagnosis was associated with a 78% higher risk of lung cancer as a second primary (SHR = 1.78; 95% CI, 1.43–2.21; **<u>Figure 5A</u>**)

**Figure 5.**
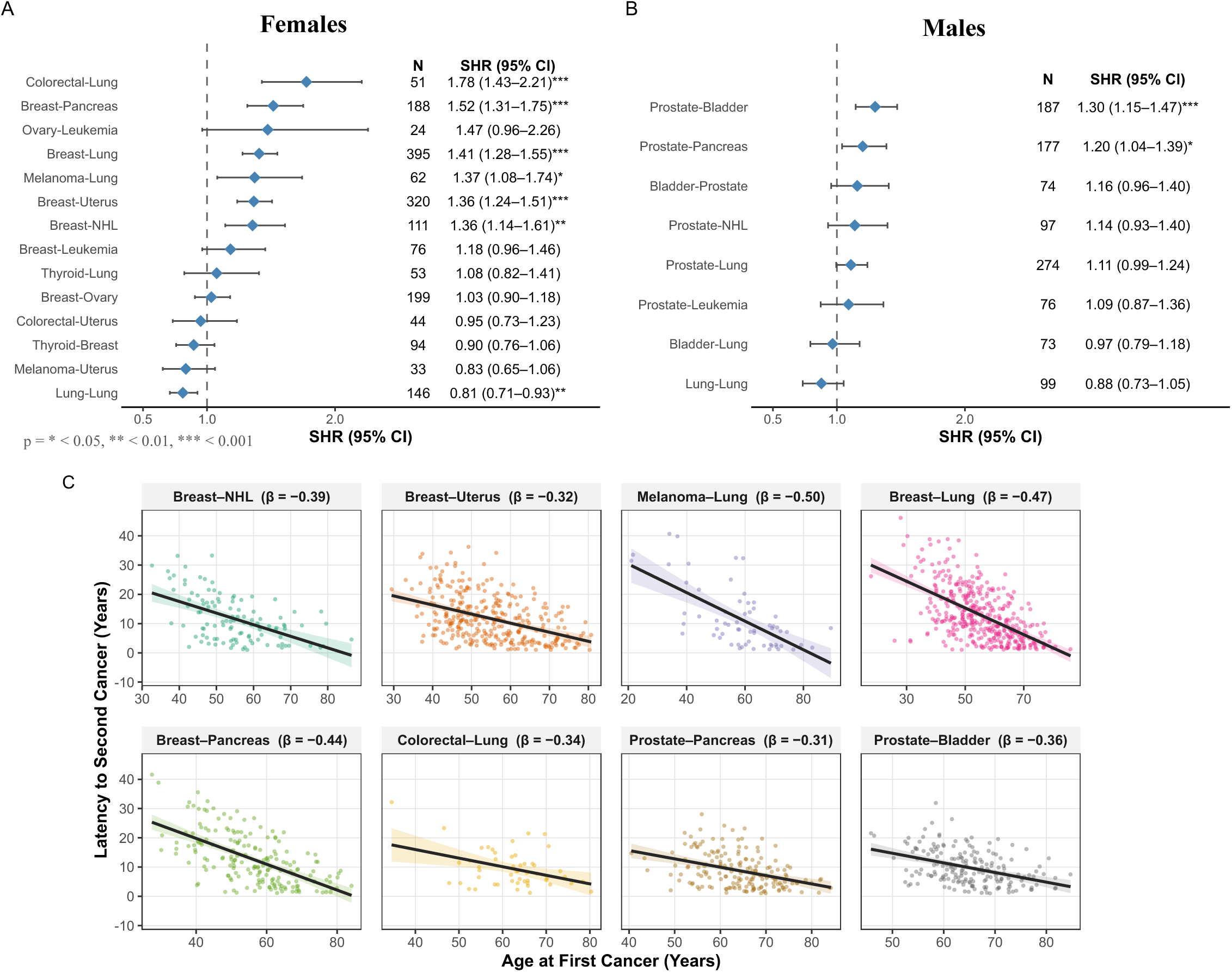
Association between older age at first cancer diagnosis and risk of second primary cancers. (A) Females and (B) Males. Points represent subdistribution hazard ratio (SHR) estimates per 1-standard deviation increase in age at first cancer diagnosis, and horizontal bars indicate 95% confidence intervals. The dashed vertical line denotes SHR = 1 (no association). N represents the number of events for each specific cancer pair. P-values are adjusted for multiple comparisons. (C) Scatterplots show the association between age at first cancer diagnosis and latency to second cancer (in years) among individuals who developed a second primary cancer, with lines showing the linear fits with 95% confidence bands. *β denotes the estimated change in latency per one-year increase in age at first cancer*.

In males, older age at first cancer diagnosis was likewise associated with an increased cumulative incidence of second primary cancers, including prostate–bladder and prostate–pancreas (**<u>Figure 5B</u>**).

Among pairs for which older age at first diagnosis was significantly associated increased risk of a second cancer, linear regression was used to descriptively examine the association between age at first cancer diagnosis and latency to second cancer (**<u>Figure 5C</u>**). Older age at first diagnosis was consistently associated with shorter time to second cancer, with estimated reductions in latency of approximately 0.3 to 0.5 years per one-year increase in age at first cancer.

As a sensitivity analysis, we restricted the sample to individuals who developed a second primary cancer and, for the significant pairs above, used logistic regression to examine whether older age at first diagnosis was associated with higher odds that the second primary was the specific cancer of interest, rather than another second-cancer type (**<u>Supplementary Figure 3</u>**). For example, in the breast–pancreas pair, older age at first (breast) cancer diagnosis was associated with increased odds of pancreatic cancer as the second primary, relative to other second primary cancers, among breast cancer patients. Results were statistically significant for most cancer pairs identified in the Fine–Gray regression analysis.

Among the 37,721 germline-consented non-synchronous patients, germline PV burden showed graded associations with excess risk in canonical hereditary cancer pairs, including breast-ovary (p < 0.001), breast-pancreas (p < 0.001), and prostate-pancreas (p = 0.038). Notably, elevated risk was not confined to PV but also observed even among germline-tested non-carriers. SIRs remained markedly elevated for breast–ovary (SIR=6.2), breast–pancreas (SIR=6.6), and prostate–pancreas (SIR=4.0), indicating substantial inherited and non-inherited contributions beyond monogenic susceptibility.

In support of this, PRS for the second cancer was associated with risk of that cancer for pancreas-related hereditary pairs (e.g. pancreatic cancer PRS predicts pancreatic cancer in breast cancer patients; breast–pancreas: OR = 1.66; 95% CI: 1.39–1.98; n = 150 events); prostate–pancreas: OR = 1.37; 95% CI: 1.13–1.68; n = 135 events), whereas the association for breast–ovary was attenuated (OR = 1.14; 95% CI: 0.97 – 1.35; n = 168 events), possibly due to the overall lower performance of ovarian PRS.

We further identified nominal support for first cancer PRS associations in second cancer risk. For the breast–pancreas cancer pair (**<u>Figure 6</u>**), breast cancer patients in the highest decile of breast cancer PRS demonstrated approximately twofold higher risk of pancreatic cancer compared with those in the lowest PRS decile (SHR = 2.22; 95% CI, 1.10–4.48; n = 150 events). This replicated when restricting to white individuals (European and Ashkenazi Jewish) (SHR = 2.10; 95% CI, 1.02–4.34; n = 126 events). Notably, individuals in the top decile of the breast cancer PRS were 57% more likely to also fall within the top decile of pancreatic cancer PRS (Fisher’s exact test; OR = 1.57; 95% CI, 1.17–2.09), despite no overlapping SNPs between the two PRS, supporting a shared polygenic basis underlying susceptibility to both cancer types. In contrast, we did not identify associations between either breast PRS and risk of ovarian cancer (breast–ovary pair) or prostate PRS and risk of pancreatic cancer (prostate–pancreas pair, **<u>Figure 6</u>),** potentially due to cancer-specific genetic relationship or limited power.

**Figure 6.**
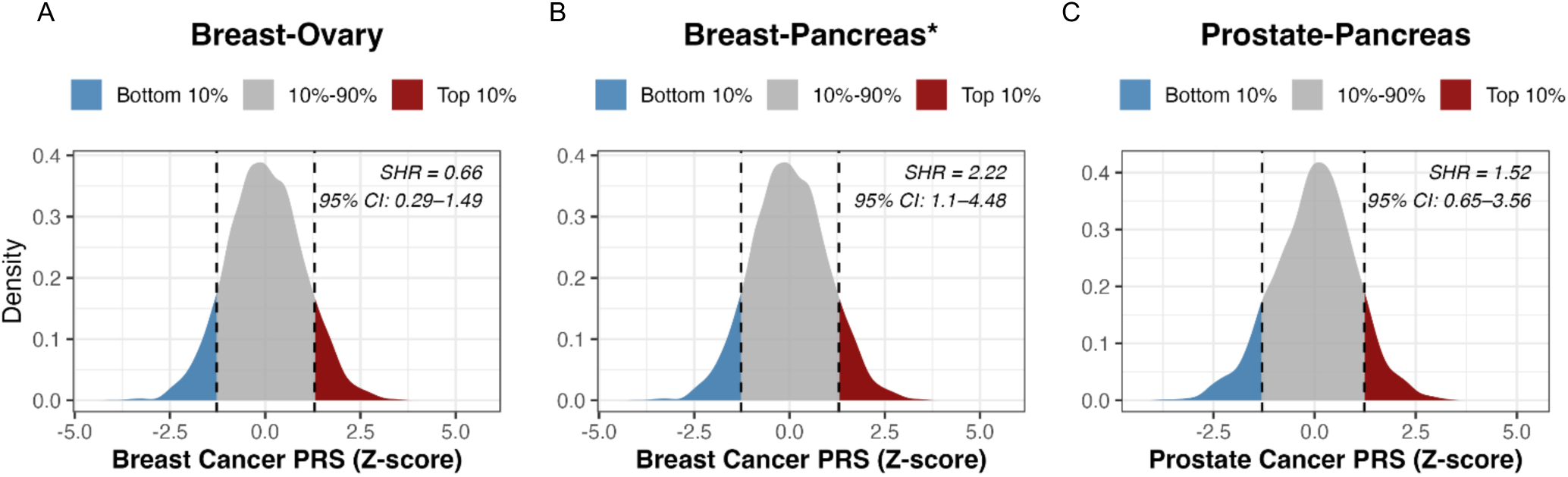
Associations between first primary cancer polygenic risk and risk of second primary cancers. Density plots represent the distribution of the first primary cancer PRS (breast or prostate) in the analyzed cohort, with shaded regions indicating the extreme deciles used for risk estimation. Subdistribution hazard ratios (SHRs) compare the risk of developing a second primary cancer between individuals in the highest versus lowest PRS deciles. A significant association was observed for the breast–pancreas pair (SHR = 2.22; 95% CI, 1.10–4.48), suggesting that individuals with higher breast cancer PRS have an increased risk of subsequent pancreatic cancer. Event counts for each cancer pair are as follows: breast–ovary (n = 168 events), breast–pancreas (n = 150 events), and prostate–pancreas (n = 135 events). *Asterisks (*) denote statistically significant associations between first-cancer PRS and risk of the corresponding second primary cancer*.

In parallel, treatment-related exposures were evaluated within our cohort (independent of SEER comparisons). In time-varying Cox regression models, tamoxifen use after breast cancer was associated with increased uterine cancer risk (HR = 2.11; 95% CI, 1.55-2.88; n = 317 events), anthracycline exposure after breast cancer was associated with increased leukemia risk (HR = 2.49; 95% CI, 1.39-4.46 n = 72 events), and PARP inhibitor exposure after ovarian cancer was associated with markedly increased leukemia risk (HR = 6.09; 95% CI, 2.53-14.68 n = 24 events). Given that treatment assignments were not randomized, these estimates represent associations rather than causal effects.

Sensitivity analyses using alternative exposure definitions yielded consistent results (**<u>Supplementary Table 4</u>**).

### Multifactorial predictive modeling and risk stratification of hereditary MPC

We next tested whether integrating clinicogenomic features can identify survivors at highest risk for metachronous second primaries in canonical hereditary cancer pairs (breast-ovary, breast-pancreas, and prostate-pancreas). For the breast–ovary and breast–pancreas cancer pairs, the cohort included 4,657 individuals with breast cancer as the first primary malignancy. Among these, the incidence of metachronous second primary ovarian cancer was 3.6% (168 events), and the incidence of second primary pancreatic cancer was 3.2% (150 events). The median time to second metachronous ovarian and pancreatic cancers was 9.8 and 10.7 years, respectively. For the prostate–pancreas cancer pair, the cohort included 3,773 individuals with prostate cancer as the first primary malignancy. Among these, 3.6% (135 events) developed a metachronous second primary pancreatic cancer, with a median time to pancreatic cancer of 6.5 years.

The selection of the LASSO penalty parameter (λ) and corresponding performance across λ values are shown in **<u>Supplementary Figure 4</u>**. Model coefficients for each cancer pair are provided in **<u>Supplementary Table 5</u>.**

The MPC-PREDICT models demonstrated strong discrimination across the different time horizons (**<u>Figure 7A-C</u>**). In the training set, AUCs ranged from 0.70 to 0.80 across the 5-, 10-, and 15-year time points. Importantly, this discriminatory performance remained consistent in the validation cohort. In validation, the breast–ovary model achieved AUCs of 0.71 (95% CI, 0.62–0.79), 0.69 (95% CI, 0.62–0.77), and 0.72 (95% CI, 0.64–0.80) at 5, 10, and 15 years from the start of follow-up, respectively. For breast–pancreas, AUCs were 0.81 (95% CI, 0.74–0.88), 0.70 (95% CI, 0.59– 0.81), and 0.71 (95% CI, 0.60–0.82), and for prostate–pancreas, AUCs were 0.70 (95% CI, 0.56– 0.85), 0.74 (95% CI, 0.63–0.85), and 0.69 (95% CI, 0.57–0.81).

**Figure 7.**
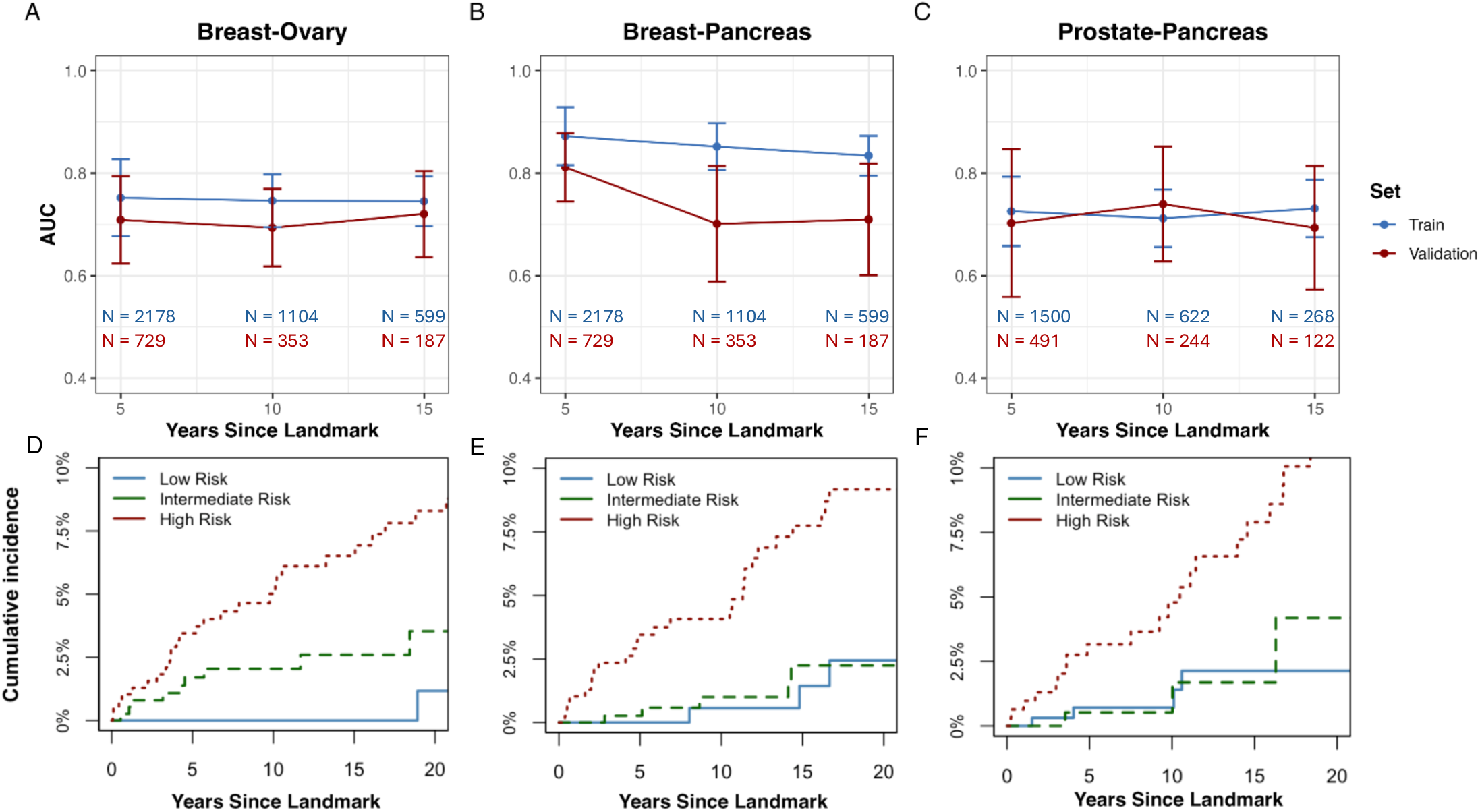
Model discrimination and risk stratification for second primary cancer prediction. (A–C) Time-dependent discrimination of the MPC-PREDICT models across cancer pairs, showing area under the receiver operating characteristic curve (AUC) at 5, 10, and 15 years from the 1-year landmark date (1-year after first cancer diagnosis) for the breast–ovary (A), breast–pancreas (B), and prostate–pancreas (C) models. Blue lines represent performance in the training set and red lines represent performance in the validation set; error bars indicate 95% confidence intervals. Discrimination was strongest for the breast–pancreas model at earlier time points and more stable performance across time for the breast–ovary model. (D–F) Risk stratification based on tertiles of the linear predictor derived from the training set, applied to the validation set. Cumulative incidence are shown for low-, intermediate-, and high-risk group1s for the breast–ovary (D), breast–pancreas (E), and prostate–pancreas (F) cancer pairs. Clear separation of curves was observed across all cancer pairs indicating effective risk stratification. *N denotes the number of individuals under observation (at risk) at each time point*.

For the breast–ovary cancer pair, the C-index was 0.76 (95% CI, 0.72–0.81) in the training set and 0.74 (95% CI, 0.67–0.81) in the validation set. For breast–pancreas, the C-index was 0.85 (95% CI, 0.81–0.89) in the training set and 0.77 (95% CI, 0.71–0.84) in the validation set. For prostate–pancreas, the C-index was 0.73 (95% CI, 0.67–0.78) in the training set and 0.72 (95% CI, 0.61–0.83) in the validation set.

Discriminatory ability was further supported by risk stratification using tertiles of the risk prediction scores derived from the training set. In the independent validation set, cumulative incidence of metachronous second primary cancers differed by approximately 4- to 7-fold between the highest- and lowest-risk tertiles across all hereditary cancer pairs **(<u>Figure 7D-F</u>).**

Model calibration was evaluated by comparing predicted and observed risks of second primary cancers across four risk strata, with O/E ratios across time points reported in **<u>Supplementary Figure 5</u>**. In the validation set, O/E ratios at 15 years were 0.94 (breast–ovary), 1.48 (breast–pancreas), and 0.85 (prostate–pancreas), reflecting good agreement for breast–ovary and prostate–pancreas but underestimation for breast–pancreas.

The clinical relevance of the MPC-PREDICT models for cancer prevention was further evaluated using 15-year decision curve analysis (DCA) (**<u>Supplementary Figure 6</u>**). Across all cancer pairs, the models demonstrated consistently higher net benefit than both *surveil-all* and *surveil-none* strategies, suggesting the models can help identify individuals meriting extended surveillance for second primary cancers, while reducing unnecessary surveillance for low-risk patients. Notably, 15-year absolute risk was generally below 10% (**<u>Figure 7D–F</u>**), consistent with the rarity of the outcome and supporting the focus on lower, relevant threshold probabilities.

Across hereditary cancer pairs, high-risk groups demonstrated distinct demographic, ancestry, and genetic profiles compared with intermediate- and low-risk groups. Individuals in the high-risk tertile were older at their first cancer diagnosis than those in the intermediate- and low-risk groups, with mean differences of 8 years for breast–ovary, 12 years for breast–pancreas, and 3 years for prostate–pancreas. In the breast–ovary cohort, *BRCA1* pathogenic variants were more prevalent in the high-risk group (9%) compared to the intermediate- and low-risk groups (1%). Similarly, in the breast–pancreas cohort, *BRCA2* pathogenic variants were more frequent in the high-risk group (6%) than in the intermediate- and low-risk groups (2%). In the breast–pancreas cohort, individuals in the high-risk group were less likely to be Black/African American (3% vs. 12%) and more likely to be of Ashkenazi Jewish ancestry (24% vs. 10%). Notably, PRS for the target second cancers (ovary and pancreas) were significantly elevated within their respective high-risk groups (Wilcoxon nominal p < 0.001), confirming that these scores track increased risk of the target second cancers. Beyond cancer-specific risk, we observed cross-cancer pleiotropic signals. In addition to the previously noted breast cancer PRS association in the breast–pancreas pair **(<u>Figure 6</u>**), prostate cancer PRS was elevated in the breast-pancreas high-risk tertile (Wilcoxon nominal p = 0.003), while testicular cancer PRS was elevated in the prostate–pancreas high-risk tertile (Wilcoxon nominal p < 0.001), despite neither corresponding to the index or target cancer in those pairings. Together, these findings suggest that second cancer risk is driven by both clinical and genetic factors, including cancer-specific and pleiotropic genetic susceptibility across cancer types.

Because rare monogenic cancer susceptibility genes are well-established risk factors and emerged among the influential features in MPC-PREDICT models, PV status was evaluated as a standalone predictor across each hereditary cancer pair (**<u>Figure 8</u>**). NCCN-defined susceptibility genes for each cancer type were identified (**<u>Supplementary Materials and Methods</u>**), and patients harboring a PV in any cancer-specific gene in the cancer-pair were classified as high-risk carriers. Discriminatory performance of PV status alone was poor, approximating random chance to minimal predictive power (15-year AUC ∼ 0.53–0.61). In contrast, MPC-PREDICT models integrating clinical and genomic features achieved substantially higher discrimination (15-year AUC ∼ 0.70; **<u>Figure 7A-C</u>).** For direct comparison, ROC curves are shown at 15 years in **<u>Figure 8</u>**, while time-dependent AUCs across 5-, 10-, and 15-year horizons are presented in **<u>Supplementary Figure 7</u>** for PV-only models.

**Figure 8.**
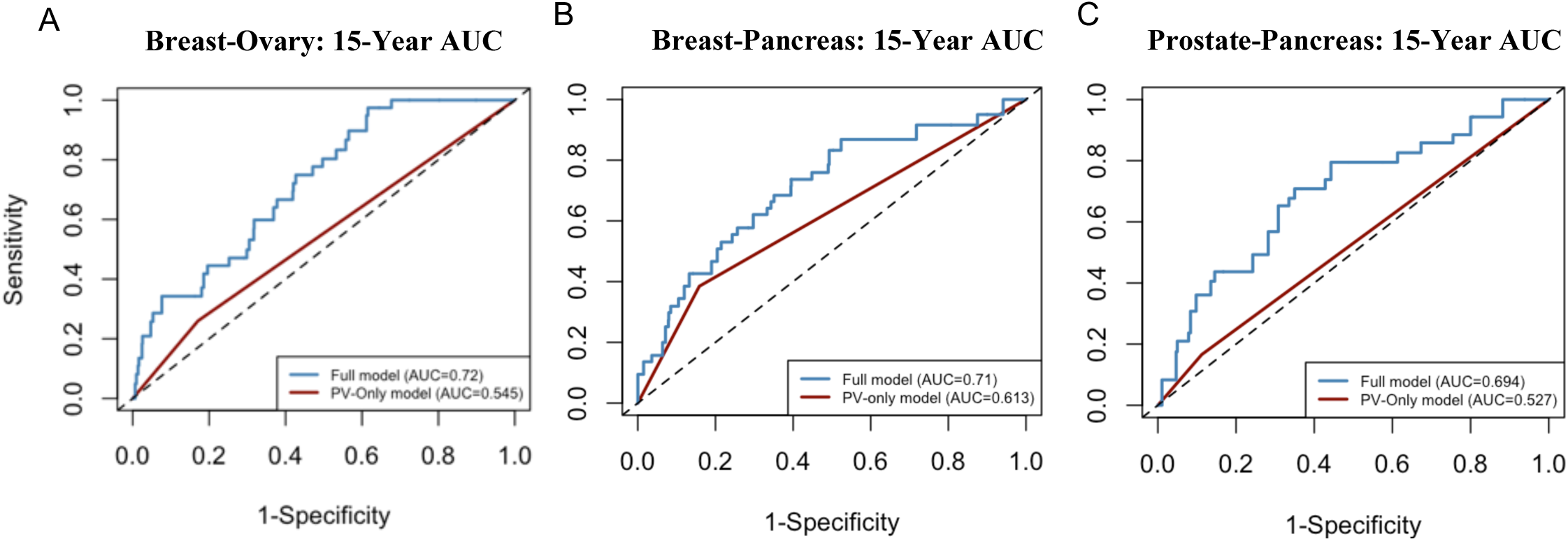
Limited discrimination of pathogenic variant status alone compared with full models. Time-dependent ROC curves at 15 years from the 1-year landmark for (A) breast–ovary, (B) breast–pancreas, and (C) prostate–pancreas cancer pairs. The full model (blue) outperforms the pathogenic variant (PV)–only model (red) across all comparisons. Dashed lines indicate no discrimination (AUC = 0.5).

A similar germline-only modeling approach was evaluated across the full cohort (including both synchronous and metachronous cancer pairs due to low sample size) for Lynch syndrome–associated colorectal–uterine cancers. Although colorectal patients carrying Lynch syndrome PV’s (*MLH1, MSH2, MSH6, PMS2, EPCAM*) demonstrated increased relative risk of second uterine cancers (SHR = 5.19; 95% CI, 2.25–12.0), discriminatory performance remained limited, with time-dependent AUCs ranging from 0.56 to 0.64 (**<u>Supplementary Figure 7</u>**), consistent with a predictor characterized by high effect size but low population prevalence.

## Discussion

This study establishes a framework for programmatic classification of MPC, enabling population-scale analysis of second cancer risk and early-stage clinicogenomic risk modeling, potentially informing future clinical tools for survivorship care. To our knowledge, this represents, among cancer pairs, the first study to combine large-scale, cancer-specific MPC classification with a multifactorial predictive model integrating germline, polygenic, clinical, and treatment data to jointly assess excess risk of distinct second primary cancers. Clinically, the findings underscore a critical public health issue: the absolute burden of metachronous primary cancers increased substantially with longer survival, as shown for breast and prostate cancer survivors. This highlights the importance of studying second primary cancer risk as cancer outcomes improve. Many of the metachronous cancer associations reported here, including breast–lung and breast–uterus, suggested in our prior analysis [15], were confirmed in a substantially larger analytical cohort of 77,060 cases. In addition, we identify additional cancer pair associations, such as breast-NHL, and prostate-leukemia, some of which were noted in meta-analyses or case series [24, 25], but are not explained by known hereditary cancer syndromes or lifestyle or treatment exposures.

We validated the fidelity of MPC phenotyping by comparing an IARC-informed classifier against expert adjudication. Since residual misclassification was concentrated in a small set of ambiguity-prone sites, and binary concordance was substantial (Cohen’s κ = 0.88), this confirms the system’s utility for real-world analyses without systematic inflation of MPC burden. Qualitatively, 22 cancer pairs demonstrated statistically significant excess risk, with 16 pairs showing greater than twofold risk, representing clinically meaningful associations. Many of these associations corroborate established clinical risk factors. For example, excess risk of second lung cancer was associated with smoking status [26,27], and obesity was a key determinant of breast–uterine cancer risk [28,29]. We also reproduced the association of increased uterine cancer risk following tamoxifen therapy for breast cancer [30], consistent with evidence suggesting tamoxifen acts as a non-genetic oncogenic driver by activating PI3K–AKT–mTOR signaling through ER-mediated *IGF1R* crosstalk, thereby phenocopying canonical driver mutations such as *PIK3CA* [31]. Additionally, we observed leukemogenic risk following anthracycline exposure [32] and identified a PARP inhibitor–associated increase in subsequent leukemia risk after ovarian cancer [33], although precise estimates were limited by low event counts.

Age at first cancer diagnosis emerged as a consistent predictor of MPC risk, serving as a proxy for cumulative biological vulnerability. Since cancer incidence increases with age due to somatic damage and declining tissue suppression of malignant clones, [34, 35] older age [36] at diagnosis was associated with a shorter interval to a second primary cancer. This implies that aging not only increases the probability of second primaries, but also accelerates their evolution, which has direct implications for surveillance in aging survivors. While age acts as a proxy for cumulative replicative opportunity [37, 38], multivariable analyses shows that inherited susceptibility and clinical exposures may shift age-related baseline risk. This analysis indicates that MPC occurrence is not purely stochastic, and subsets remain clinically stratifiable based on clinicogenomic features.

Most notably, analyses of canonical hereditary cancer pairs, including breast-ovary, breast-pancreas, and prostate-pancreas, revealed persistent high excess second cancer risk even after stratification by PV carrier status. This suggests that monogenic susceptibility alone does not fully explain second cancer risk within these pairs. One plausible explanation is that common genetic variants with small effect sizes contribute to second cancer risk, including both tumor-specific variants and pleiotropic loci shared across cancer types. Prior work has shown that cross-cancer PRS captures shared genetic architecture between different tumor types [39, 40], but whether shared genetic loci contribute to distinct MPC pairs remains underexplored. Initial modeling found suggestive evidence supporting the role of polygenic susceptibility contributing to second cancer risk beyond high-penetrance variants among certain pairs. We did note a significant association of higher breast cancer PRS and risk of subsequent pancreatic cancer; individuals in the top decile of breast cancer PRS were more likely to also reside in the top decile for pancreatic cancer PRS. Similarly, MPC-PREDICT models identified suggestive cross-cancer associations for prostate cancer PRS in the breast–pancreas pair and testicular cancer PRS in the prostate–pancreas pair. Such PRS associations warrant further exploration, as shared biological features among components in PRS may be related to DNA repair capacity, hormone and metabolic states, immune surveillance, stem-cell turnover, and the tissue or tumor microenvironment that are relevant across cancers.

Additionally, although high-penetrance genes such as *BRCA1/2, MSH2, MLH1, TP53, PTEN,* and *CDH1* remain central to cancer surveillance paradigms, models based on these rare variants alone showed limited discrimination in our analyses. Consistent with this, population attributable risk fractions (PAR) for *BRCA*-associated MPC ranged from moderate to weak, with higher estimates observed for breast–ovarian cancer (13.4% for *BRCA1*-associated ovarian cancer following breast cancer) and substantially lower values for pancreatic-associated pairs (5.7% for *BRCA2*-associated pancreatic cancer following breast cancer and 2.9% for *BRCA1*-associated pancreatic cancer following prostate cancer), reflecting the low prevalence of PV carriers despite elevated individual relative risks. Crucially, *BRCA1/2* variants were not retained in the prostate–pancreas MPC-PREDICT model, likely reflecting their limited PAR contribution in this context. In contrast, cancer-specific PRS were consistently retained with moderate-magnitude coefficients across all MPC-PREDICT models and showed higher population-level impact. For example, when defining high genetic risk as the top quartile, pancreatic cancer PRS had a PAR of 15.1% in the prostate–pancreas pair, compared to 2.9% for *BRCA1*. These findings indicate that common variant architecture contributes to identifying groups of patients at increased risk of MPC, including those traditionally attributed to monogenic hereditary syndromes, and support a continuum model of cancer susceptibility rather than a binary classification based on monogenic carrier status. Clinically useful risk stratification emerged most robustly through integrated genetic and clinical modeling, supporting a shift beyond gene-centric approaches toward data-driven survivorship surveillance. In practice, multifactorial models could better identify subsets of cancer survivors at elevated risk who may benefit from chemoprevention, interception, secondary prevention, risk-reducing surgeries, or investigational approaches including circulating cell-free DNA–based detection assays [41–43] or serial assessment of clonal hematopoiesis [44].

The model utilized here represents an application of machine learning approaches to a multi-dimensional dataset, incorporating EMR-derived clinical features not typically used in second cancer risk analyses [45]. Our framework enables generation of individualized absolute risk estimates at 5-, 10-, and 15-year time horizons, as well as stratification into cancer-specific risk groups. Consistent with prior applications of second cancer risk prediction [46], we employed Fine-Gray regression to account for competing events which are inherent to cancer survivor populations. Across cancer pairs, the models demonstrated good discrimination together with consistent evidence of clinical utility. In this setting, where no single genetic or clinical factor fully explains second cancer risk and events are rare, multivariable approaches grounded in conservative risk modeling are well suited, as reflected by the models’ AUC and ∼4–7-fold separation in cumulative incidence between low- and high-risk groups across hereditary cancer pairs. DCA further supported these findings, demonstrating improved net benefit over *surveil-all* and *surveil-none* strategies across clinically meaningful thresholds. These results are consistent with prior work in contralateral breast cancer risk prediction [46], indicating comparable clinical utility despite differences in cancer types and modeling approaches.

Despite the model’s strengths, several limitations must be acknowledged. We excluded synchronous cancers, limiting conclusions about shared etiologies, and the analyses were conducted within a single tertiary-care sequencing cohort, restricting generalizability due to potential referral biases and detection of incidental findings at time of cancer diagnosis [47]. In addition, a substantial proportion of individuals with multiple primary cancers underwent T:N sequencing after the diagnosis of a second cancer, which may introduce some bias in inference analyses and affect the precision of effect size estimates. External validation in large, diverse, population-based cohorts is essential. However, high quality data spanning years of medical care may be difficult to obtain despite the increasing availability of the electronic medical and billing records [48, 49]. While such comprehensive datasets are currently limited, ongoing improvements in large-scale clinical NLP-based extraction from EMR [8], together with the growing population of cancer survivors, are expected to enable these analyses in the near future. Our approach provides a unified competing-risk framework for cancer pair–specific risk prediction across hereditary cancer combinations, whereas prior studies often estimate second cancer risk without distinguishing tumor pairings or focus on a single combination such as contralateral breast cancer [46, 50, 51].

These findings underscore the need to elucidate the biological mechanisms underlying gene–environment interactions, epistatic effects, and other endogenous and exogenous factors associated with second primary cancer risk. Future approaches could pretrain a multimodal foundation model on longitudinal clinicogenomic data, integrating event histories, laboratory measurements, and therapies to generate calibrated, time-resolved second cancer risk predictions. Finally, MPC-PREDICT is the first set of models to integrate genomic, environmental, and lifestyle factors for prediction of hereditary second cancers. This work provides an adaptable framework for future clinical tools to support prevention, surveillance, and risk-informed decision-making among cancer survivors and high-risk individuals.

## Acknowledgements

This work was supported by the MSK Support Grant/Core Grant (P30CA008748), the MSK Molecular Diagnostics Service in the Department of Pathology, the Marie-Josée and Henry R. Kravis Center for Molecular Oncology, the Movember–Prostate Cancer Challenge Award, the Prostate Cancer Foundation (Challenge Award 18CHAL05), the National Cancer Institute (P01CA228696, P30CA008748, U01CA257679), the Robert and Kate Niehaus Center for Inherited Cancer Genomics and the Andrew Sabin Family Foundation, the CureBRCA Foundation, and the Breast Cancer Research Foundation. The authors also acknowledge The Clinical Genetics Service, Department of Medicine, Memorial Sloan Kettering Cancer Center. The funders had no role in study design; data collection, analysis, or interpretation; manuscript preparation; or the decision to submit for publication.

## Disclosures of professional relationships and financial interests

***Y.L.L*** reports : Artios Pharma Limited, AstraZeneca, Curio Science LLC, GlaxoSmithKline, HMP Oncology Learning Network, MDoutlook, LLC, Myriad Genetics, Repare Therapeutics, Topline Bio, Total Health Conferencing, Gilead Sciences, AbbVie; ***J.J:*** MDSeq Inc; ***S.M:*** Pfizer, Inc., Regeneron Pharmaceuticals, Inc.; ***M.F.Berger:*** AstraZeneca, JCO Precision Oncology, Journal of Molecular Diagnostics, Paige.AI, Inc., SOPHiA GENETICS S.A.; ***D.S:*** Antares Therapeutics, Inc., BridgeBio Inc., Corramedical, Inc., Elsie Biotechnologies, Inc., Fog Pharmaceuticals, Inc., Fore Biotherapeutics, Function Oncology, Inc., Meliora Therapeutics, Paige.AI, Inc., Pfizer, Inc., Pyramid Biosciences, Inc., Scorpion Therapeutics, Inc.; ***Z.S:*** American Society of Clinical Oncology (ASCO), SOPHiA GENETICS S.A., UptoDate; ***N.S:*** Innovation in Cancer Informatics, Stand Up to Cancer. All other authors report no conflicts of interest. ***D.M:*** AstraZeneca, **G.S:** AbbVie, Lilly USA, LLC, BeiGene USA, Inc., Medscape, Bristol-Myers Squibb, ModeX Therapeutics Inc., Ellipses Pharma, Foresight Diagnostics, Inc., Pfizer, Inc., Genentech, Research to Practice, GenMab, Roche, Gilead Sciences, Inc., Scientific Education Support Ltd., Ipsen Pharma, SERB Pharmaceuticals, Janssen Global Services, LLC, Treeline Biosciences, Inc.

## Author Contributions

J.A, K.O, V.J conceived of the presented idea. J.A, I.O, A.M developed the theory and performed the computations. J.A, I.O, A.M, V.R, M.C, Y.L.L, K.O, V.J, verified the analytical methods. J.A, A.M, Y.K, T.P, J.C, C.F, R.K, S.M, M.C, E.K, J.V, I.O, helped with generation of data and data wrangling. J.A, K.O, V.J, wrote the first draft of the manuscript. J.A, I.O, A.M, Y.L.L, T.P, V.R, J.J, M.C, A.K, Y.K, E.K, S.M, A.L, L.B, R.K, S.M, C.F, M.B, C.B, J.B, V.S, G.S, D.M, M.B, D.S, Z.S, N.S, J.Z, K.O, V.J read and edited the manuscript. K.O and V.J supervised this work. All authors provided critical feedback and helped shape the research analysis presented in the final manuscript.

