## Supplementary Materials and Methods for "Integrated Clinicogenomic Risk Modeling for Metachronous Second Primary Cancers"

**^1^Department of Medicine, Memorial Sloan Kettering Cancer Center, New York, NY, USA.**

**^2^Department of Biostatistics and Epidemiology, MSKCC, New York, NY, USA**

**^3^Center for Molecular Oncology, MSKCC,** **New York, NY, USA**

**^4^Department of Pathology, MSKCC, New York, NY, USA**

**^5^Weill Cornell Medical College, New York, New York.**

**^6^Neihaus Center For Inherited Cancer Genomics, MSKCC, New York, NY, USA**

† Lead contact author

**MPC Algorithm Design:**

In this section we expand upon the steps involved in building the multiple primary cancers (MPC) classifier. Each tumor record was preprocessed using standardized ICD-O-3 topology (C00.0–C96.9) and morphology (M8000–M9999/x) codes [1]. Tumors lacking essential fields (topology, or morphology) were excluded. Only invasive malignancies (behavior code /3) were retained, with the exception of ductal carcinoma in situ (DCIS) of the breast (/2), which was included due to its clinical relevance, because it is an actionable premalignancy with a relatively high risk of progression [2] and because retaining DCIS cases preserved breast cancer sample sizes for downstream statistical inference without discarding clinically meaningful events.

Subsequently, cancer types were further standardized and grouped according to **OncoTree,** MSKCC’s hierarchical cancer ontology [3], which harmonizes tumor classification information into clinically relevant disease categories.

Tumors were classified at the patient level using longitudinal tumor registry data. The algorithm primarily follows the International Agency for Research on Cancer’s (IARC) *International Rules for Multiple Primary Cancers* [4], with additional exception handling based on clinical expertise from oncologists at MSKCC, as well as supplemental rules inspired by and adapted from Asia-Pacific Journal of Cancer Prevention (APJCP) guidelines [5].

Core IARC-Based Rules:

(i) Morphology grouping: Tumors were grouped into broad histologic families (e.g., adenocarcinoma, squamous cell carcinoma), following the hierarchical groupings defined in IARC Table 2. Non-specific morphology codes were retained as distinct morphology groups rather than being resolved to a more specific histologic group.

(ii) Topography grouping: Related anatomical subsites were consolidated to prevent oversplitting due to minor subsite variation. In accordance with IARC guidelines, skin and colorectal subsite tumors were keyed on the full ICD-O-3 topology code rather than the consolidated group. This reflects the distinct skin anatomical locations (e.g., scalp vs. trunk), representing independent primaries, and colorectal subsites that carry distinct clinical significance that would be obscured by grouping.

(iii) Paired organs: For paired organs (e.g., breast, lung), laterality was used to distinguish independent primaries. Specifically, tumors sharing the same organ and histologic family were retained as distinct primaries only if they carried a different laterality code. Tumors with a laterality code already represented in the patient's record were deduplicated. When laterality information was unavailable for one or both tumors, the pair was deduplicated to ensure consistent stricter classification than inflation of MPC. Paired organs were therefore excluded from most downstream analyses to avoid introducing ambiguity into primary count estimates.

(iv) Systemic malignancies: Hematologic and lymphoid cancers were treated as systemic

diseases and deduplicated by morphology alone.

APJCP-Inspired Exceptions:

To address common registry ambiguities, additional rules were applied:

(i) Genitourinary tumors: Synchronous prostate and bladder adenocarcinomas diagnosed on the same date were classified as a single primary, with the bladder tumor removed when a prostate primary was present.

(ii) Colorectal tumors: Colon and rectal tumors with identical morphology and diagnosis dates were evaluated by stage; the lower-stage tumor was removed to account for tumors spanning the rectosigmoid junction.

MSKCC-Specific Rules:

(1) Temporal separation: Tumors of the same site and morphology diagnosed more than 15 years apart were classified as distinct primaries. This threshold reflects prior pan-cancer analyses showing that second primary incidence peaks approximately 10–15 years after initial cancer diagnosis [6] , with the upper bound chosen as a conservative criterion. To avoid over-splitting in patients with long follow-up, only one such time-based exception was permitted per patient.

(2) DCIS exception: Breast DCIS was retained as a primary malignancy despite its in-situ classification.

**MPC Algorithm Evaluation:**

The outputs of the MPC classifier were evaluated against a manually curated subset of 25,482 patients, all with single or multiple primary cancers, as described in [7]. Two evaluation tasks were defined:

(1) **Binary classification**, assessing whether the classifier correctly identified each patient as having a single primary cancer (SP) or multiple distinct primary cancers (MPC), relative to the expert-adjudicated *“truth set”.* Patients for whom the classifier assigned no primary cancers but had a cancer diagnosis in the *“truth set”* were retained in the denominator and counted as misclassifications.

(2) **Ordinal MPC tier concordance**, assessing agreement with the adjudicated number of distinct primary malignancies among patients classified as MPC. MPC tiers were defined based on the number of primary cancers: Tier 2 (two primaries), Tier 3 (three primaries), Tier 4 (four primaries), continuing up to Tier 7 (seven primaries; the maximum observed). These tiers enabled evaluation of algorithm performance across increasing levels of clinical complexity, as higher-order malignancy histories (e.g., ≥4 primaries) present greater challenges for automated classification.

Performance for both tasks was evaluated using accuracy (the overall proportion of correctly classified patients) and F1 score. For the binary SP versus MPC classification, sensitivity (true positive rate for MPC) and specificity (true negative rate for SP) were additionally reported among patients assigned at least one primary cancer by the classifier. Agreement beyond chance was assessed using Cohen’s κ [8], while weighted Cohen’s κ (Fleiss–Cohen weights) was used to evaluate concordance across ordinal MPC tiers [9].

**Standardized Incidence Ratios (SIR):**

SIR were generated following a previously described approach [7], defined as the ratio of observed to expected cancer counts (SIR = O/E). Expected case counts were calculated by applying SEER-21 incidence rates to accrued person-time, incorporating latency weighting (ρ = 0.9) [7]. Confidence intervals were estimated using nonparametric bootstrapping (5,000 resamples), with 95% confidence intervals defined by the empirical 2.5th and 97.5th percentiles. SIR analyses were restricted to second primary cancers only; among patients with three or more primary malignancies, only the first two diagnoses were considered to maintain a consistent at-risk definition and to avoid dependence introduced by later events. Cancer-pair combinations with sparse data (weighted expected <1 and weighted observed <2) were excluded to reduce estimate instability in the primary SIR analysis.

As a sensitivity analysis, SIR were also estimated without latency weighting (ρ = 1.0) to assess the robustness of excess risk estimates (**Supplementary Tables 1 and 2**).

**Risk Stratification by Clinical and Genetic Factors:**

This section describes the clinicogenomic factors used to stratify SIR and evaluate differential risk of site-specific second primary cancers.

*Smoking:* For cancer pairs in which lung cancer was the second primary, smoking status was analyzed to stratify excess risk. Smoking status was categorized as never versus former/current smoker based on clinical records. Stratum-specific SIR were estimated to assess differences in second primary cancer risk by smoking exposure, and excess risk was compared between strata.

*Body mass index* (BMI): BMI was classified using standard World Health Organization categories: normal weight (18.5-24.9 kg/m²), overweight (25.0-29.9 kg/m²), and obese (≥30.0 kg/m²) [10], based on the earliest available BMI measurement after first diagnosis. SIR were estimated within each BMI category, and pre-specified statistical comparisons were performed between obese and normal-weight patients.

*Germline PLP Variants:* For cancer pairs with established hereditary predisposition (breast-ovary, breast-pancreas, prostate-pancreas), germline genetic risk was stratified using NCCN-defined cancer susceptibility genes based on the presence of pathogenic or likely pathogenic (PLP) variants. Patients were classified as high risk (PLP variants in high-penetrance genes), moderate/low risk (variants in other genes included on the germline testing panel), or none (negative germline testing). Among patients consenting to clinical receipt of germline information, variant calling and assertions were made by the clinical laboratory at MSKCC, as previously described [11, 12]. Analyses utilizing germline PLP variants were restricted to patients who consented to receipt of germline testing (Figure 1). Stratum-specific SIR were estimated, and statistical comparisons were performed between high-risk carriers and non-carriers (germline pathogenic variants classification is provided below).

In parallel, polygenic risk scores (PRS) were constructed using 607 curated germline single nucleotide polymorphisms (SNPs) (**Supplementary Table 6**) derived from published instruments, with imputed genotype data generated from tumor–normal sequencing using GLIMPSE2 [13]. Cancer-specific PRS were calculated using established SNP weights from Graff et al. [14], except for breast cancer, which used weights from Mavaddat et al. [15]. To assess shared genetic predisposition across established hereditary pairs, PRS for the first primary cancer (breast or prostate) were used to stratify individuals by deciles, and Fine–Gray regression was applied to estimate the risk of second primary cancers (ovary or pancreas), comparing the highest versus lowest deciles. Models were adjusted for age at first diagnosis, BRCA1/2 status, and genetic ancestry (principal components 1–4).

*Age at First Cancer:* To evaluate age-at-onset effects, age at first cancer diagnosis was analyzed both categorically and continuously. For categorical analyses, patients were grouped into quintiles and SIR were compared between the highest and lowest quintiles (80th vs 20th percentile; late- vs early-onset group), with differences in latency to second cancer assessed using the Mann–Whitney U test. To avoid arbitrary thresholds, Fine–Gray regression [16] was used to estimate the hazard of second primary cancer per one–standard deviation increase in age, with adjustment for ancestry, BMI, smoking status, and stratification by sex. Multiple testing correction was performed using the Benjamini–Hochberg false discovery rate [17]. As a sensitivity analysis, case-only logistic regression was conducted among individuals with ≥2 primary cancers. This approach allowed us to evaluate whether age at first cancer influenced the relative distribution of specific second cancer types among individuals who developed MPC, rather than reflecting a general association between age and overall MPC risk.

Linear regression was additionally used to estimate differences in latency to second cancer (years) per one-year increase in age at first cancer diagnosis, with models adjusted for ancestry, BMI, and smoking status and restricted to cancer pairs demonstrating significant associations.

*Treatment Exposure:* Treatment exposures were modeled as time-dependent covariates, with patients contributing unexposed person-time until treatment initiation and exposed person-time thereafter, thereby accounting for treatment timing and avoiding immortal time bias. Analyses focused on predefined treatment-outcome pairs, including Poly(ADP-ribose) polymerase (PARP) inhibitor exposure and subsequent leukemia following ovarian cancer, anthracycline exposure and subsequent leukemia following breast cancer, and tamoxifen exposure and subsequent uterine cancer following breast cancer. Hazard ratios (HR) and 95% confidence intervals were estimated for each association. All analyses were adjusted for age at first cancer diagnosis, ancestry, BMI, prior radiation exposure, and prior treatment outside MSK (when available). To evaluate the robustness of treatment-associated risk estimates, sensitivity analyses were performed using alternative exposure definitions detailed in **Supplementary Table 4**. For predictive modeling using MPC-PREDICT, treatment exposures were recorded only if administered within 1 year of the first cancer diagnosis. Radiation therapy exposure was later incorporated into the MPC-PREDICT model using the same time window.

Detailed results for the SIR risk-stratification analyses are presented in **Supplementary Tables 1 and 2**.

**Germline Pathogenic Variants Classification:**

Cancer-specific susceptibility genes were defined according to National Comprehensive Cancer Network (NCCN) guidelines and used for germline-stratified SIR analyses [18, 19]. Only pathogenic or likely pathogenic variants in the genes listed below were considered highly penetrant for the specific cancer pairs evaluated in the SIR analyses. PLP variants in other genes included on the germline testing panel were classified as moderate/low penetrance, and patients with negative germline testing were classified as non-carriers. The following gene sets were applied:

(1) Breast cancer: *BARD1, BRCA1, BRCA2, CDH1, CHEK2, PALB2, PTEN, TP53, STK11, NF1.*

(2) Ovarian cancer: *BRCA1, BRCA2, ATM, BRIP1, PALB2, RAD51C, RAD51D, MLH1, MSH2, MSH6, EPCAM.*

(3) Prostate cancer: *BRCA1, BRCA2, HOXB13, ATM, TP53, CHEK2.*

(4) Pancreatic cancer: *ATM, BRCA1, BRCA2, CDKN2A, STK11, TP53, MLH1, MSH2, MSH6, PMS2.*

**MPC-PREDICT Evaluation:**

This section outlines the evaluation methods and metrics used to assess the three MPC-PREDICT models.

To select the LASSO penalty parameter (λ), each model was assessed using a 15-year time-dependent area under the receiver operating characteristic curve (ROC AUC). The optimal λ was chosen using the 1-standard error rule based on AUC performance across all iterations to favor a sparser model [20]. Final models were then refit on the full training set using the selected λ and evaluated on the independent validation set.

Model discrimination was evaluated in the validation set using time-dependent AUC at 5-, 10-, and 15-year time points from the start of follow-up, as well as concordance indices (C-index). Both metrics range from 0 to 1, with higher values indicating better discrimination. While the AUC assesses discrimination at specific time points, the C-index summarizes the model’s ability to rank individuals by risk over the entire follow-up period. Complementary to these metrics, patients were stratified into tertiles of predicted risk, and absolute risk curves were compared across strata to evaluate the model’s ability to separate clinically distinct risk groups.

Model calibration was assessed by comparing predicted and observed risks of second hereditary primary cancers across quartiles of predicted risk at each time point, with observed risks estimated using cumulative incidence functions to account for competing risks. Calibration was evaluated using the observed-to-expected (O/E) ratio. An O/E ratio greater than 1 indicates that the model underestimates risk, whereas a ratio less than 1 indicates overestimation.

The clinical relevance of the MPC-PREDICT models was evaluated using decision curve analysis [21], which quantifies the net clinical benefit of a prediction model across a range of threshold probabilities. This framework allows comparison of the model against *surveil-all* and *surveil-none* strategies, representing default approaches of intervening in all or no patients, respectively. Each threshold probability reflects the tradeoff between the benefits of identifying true cases and the harms of unnecessary intervention.
