## Supplementary Figures for "Integrated Clinicogenomic Risk Modeling for Metachronous Second Primary Cancers"

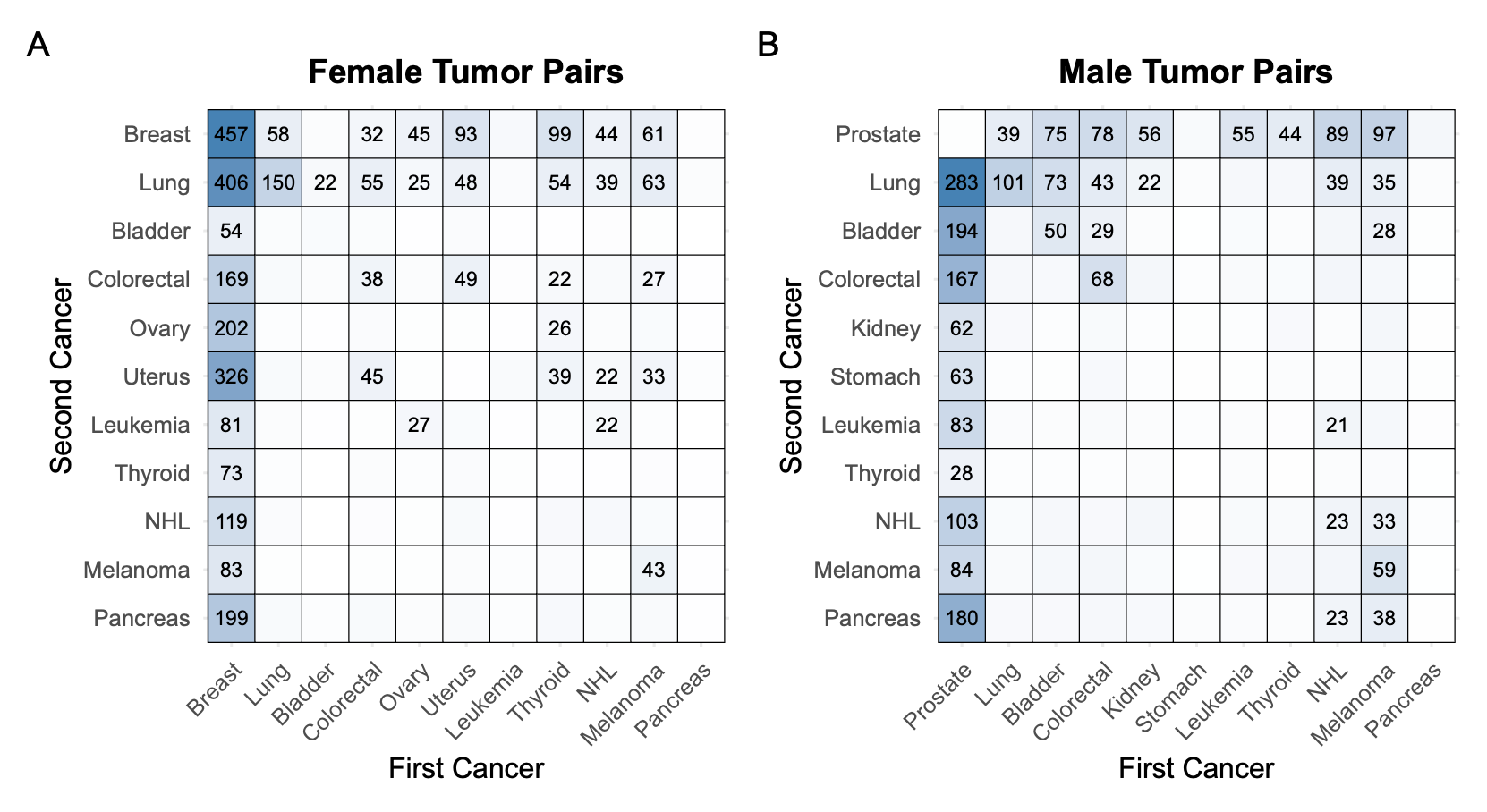

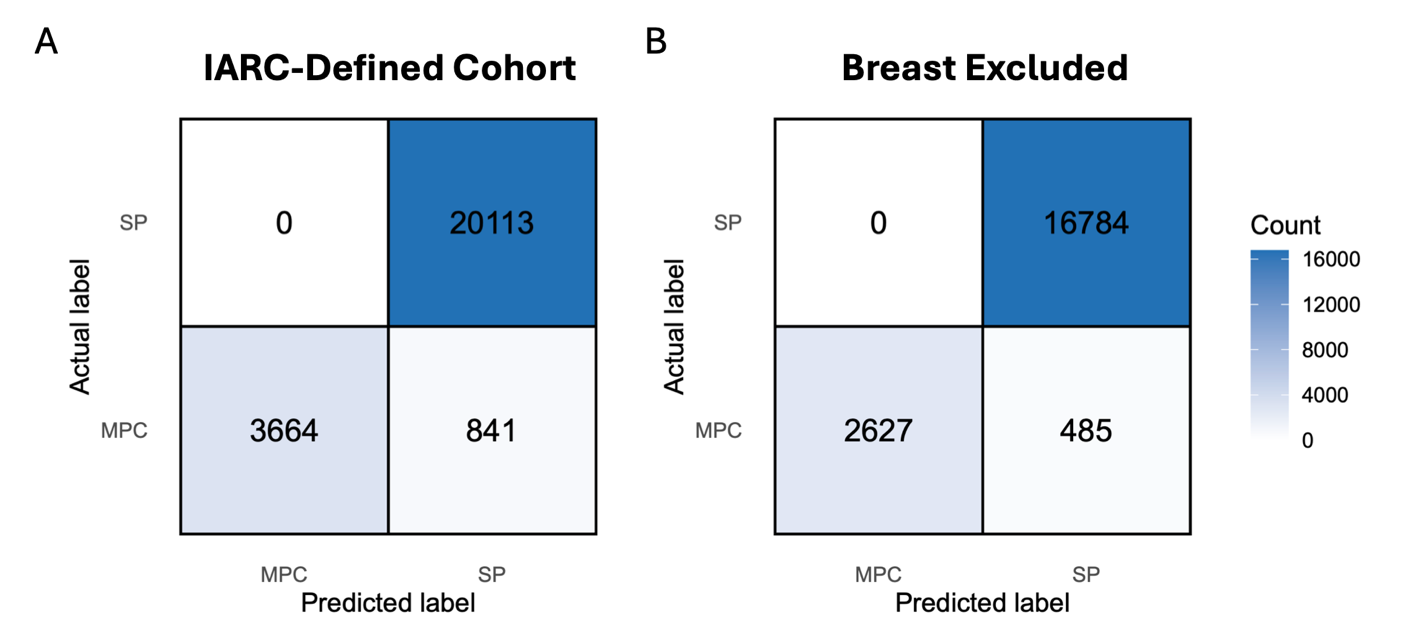

**Supplementary Figure 1. Pairwise distribution of metachronous primary cancers pairs by sex.** Heatmaps show the frequency of metachronous multiple primary cancer patients (limited to first two primaries), plotted by the site of the first cancer on the horizontal axis and the site of the second cancer on the vertical axis. Each cell indicates the number of patients diagnosed with the corresponding ordered pair; numbers are displayed in cells with >20 patients. (A) Female patients; (B) male patients. Only the most common cancer pairs in each sex are displayed. The asymmetry across the diagonal reflects the directionality of diagnosis (e.g., breast→lung vs. lung→breast). *Abbreviations: NHL:Non-Hodgkin Lymphoma.*

**Supplementary Figure 2.** **Confusion matrices for binary MPC versus SP classification.** (A) *“Truth Set”* restricted to IARC-defined primary tumors (excluding the 864 non-IARC patients classified as having no primary cancers by the MPC classifier; n = 24,618), demonstrating sensitivity of 81.4%, specificity of 100%, F1-score of 0.90. The classifier correctly identified all SP cases (0 false positives) while conservatively misclassifying a subset of MPC patients as SP (n = 841), reflecting its design preference to avoid inflating MPC counts when tumor independence is uncertain. (B) Cohort excluding breast cancer diagnoses (n = 19,896), where incomplete laterality data are a primary driver of discordance. Removing breast cases improved sensitivity to 84.5%, with specificity remaining at 100%, F1-score of 0.92, confirming that breast-related ambiguity accounts for a share of residual misclassification.

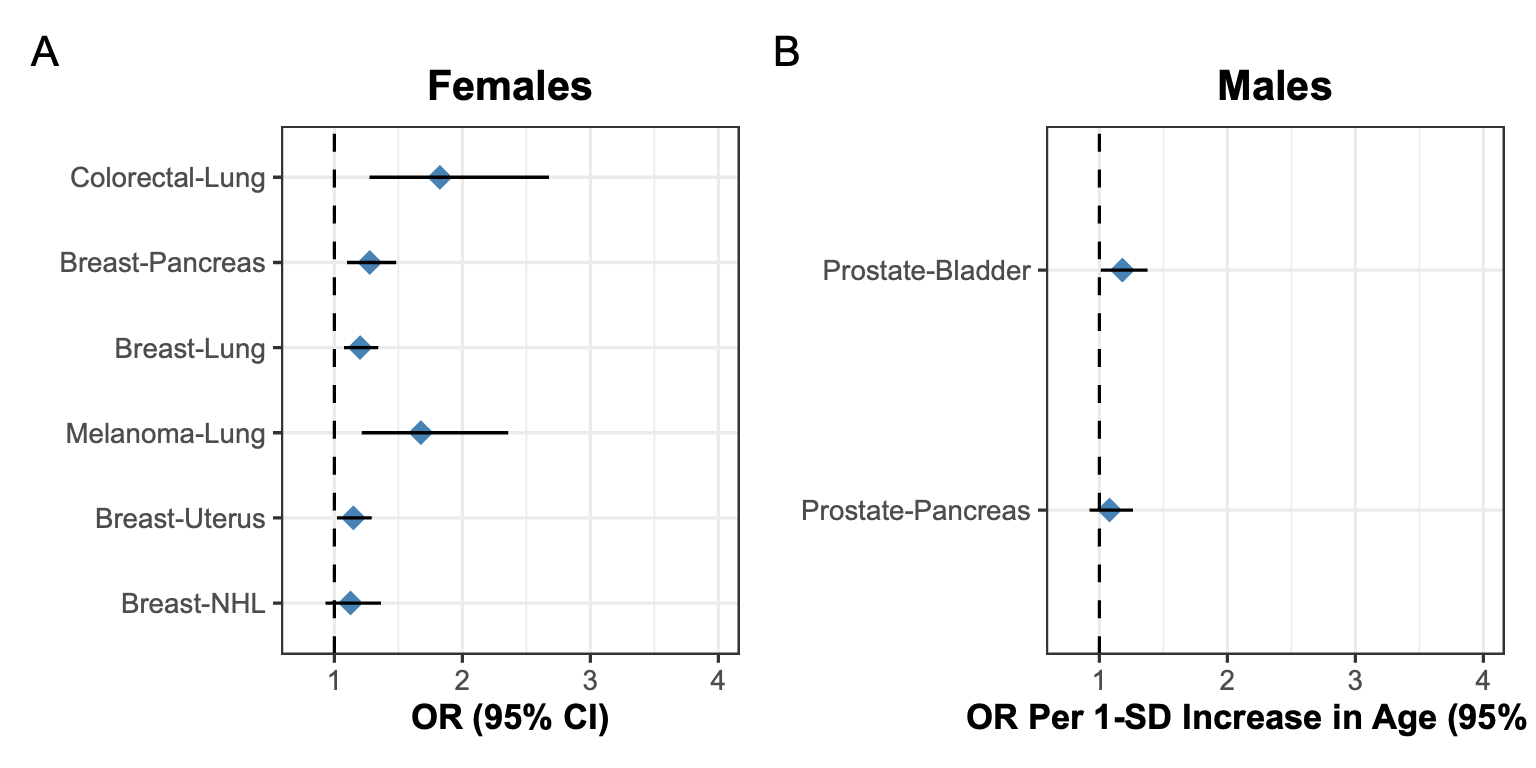

**Supplementary Figure 3. Case-case sensitivity analysis of age at first cancer diagnosis and risk and of second primary cancers.** (A) Females and (B) Males. Points represent odds ratios (ORs) per 1-standard deviation increase in age at first cancer diagnosis, with horizontal bars indicating 95% confidence intervals. The dashed vertical line denotes OR = 1 (no association). Analyses were restricted to individuals who developed a second primary cancer; thus, ORs represent the relative odds of developing a specific second cancer type compared with other second primary cancers. For example, among breast cancer survivors who developed a second primary malignancy, each 1-standard deviation increase in age at first diagnosis was associated with higher odds that the second cancer was pancreatic cancer rather than another cancer type. In females, older age at first cancer diagnosis was associated with higher odds of colorectal–lung (OR = 1.82; 95% CI, 1.27–2.68), breast–pancreas (OR = 1.28; 95% CI, 1.10–1.48), breast–lung (OR = 1.20; 95% CI, 1.07–1.34), melanoma–lung (OR = 1.67; 95% CI, 1.21–2.36), and breast–uterus (OR = 1.15; 95% CI, 1.02–1.29). In males, older age at first cancer diagnosis was associated with higher odds of prostate–bladder (OR = 1.18; 95% CI, 1.01–1.38). Breast–NHL (OR = 1.13; 95% CI, 0.93–1.36; Females) and prostate–pancreas (OR = 1.08; 95% CI, 0.92–1.26; Males) were not statistically significant in this sensitivity analysis. *N represents the number of events for each specific cancer pair.*

n = 51

n = 188

n = 395

n = 62

n = 320

n = 111

n = 187

n = 177

**OR Per 1-SD Increase in Age (95% CI)**

**OR Per 1-SD Increase in Age (95% CI)**

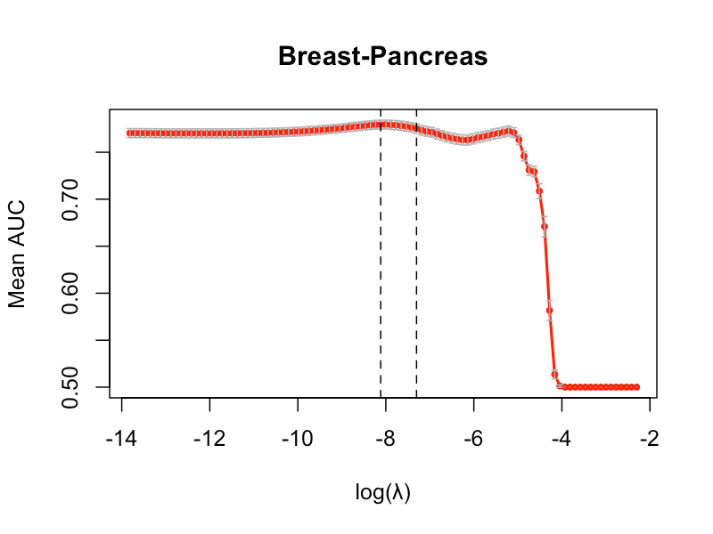

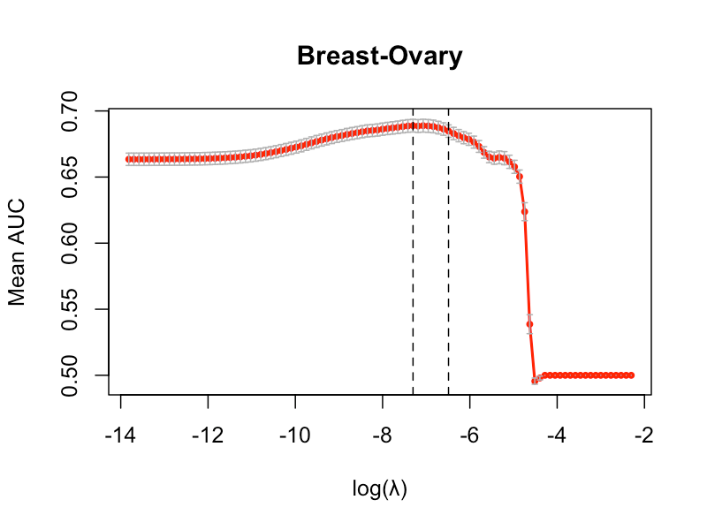

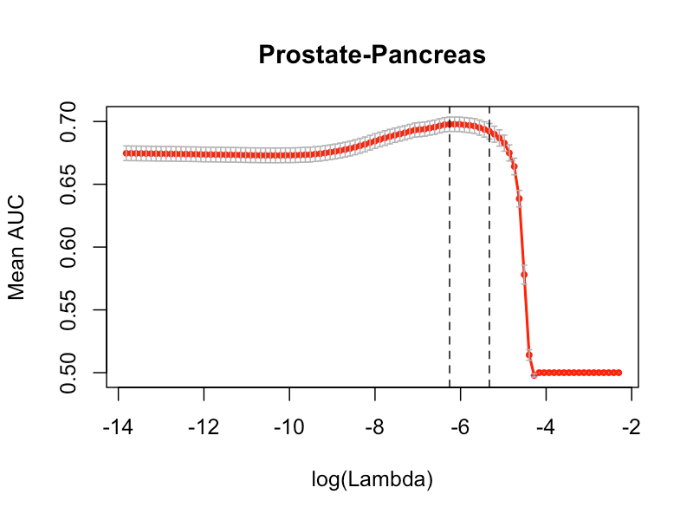

**Supplementary Figure 4. Cross-validated performance of MPC-PREDICT models across penalty parameters (λ).** Mean time-dependent AUC at 15 years from the 1-year landmark date (1 year after first cancer diagnosis) is shown across 100 Monte Carlo cross-validation iterations, with error bars indicating standard error. Vertical dashed lines indicate the selected λ values corresponding to the maximum AUC and the 1-SE rule (λ_1se), which achieved similar discrimination with fewer features. λ was selected to maximize cross-validated time-dependent AUC, aligning model selection with the primary objective of discrimination.

C

B

A

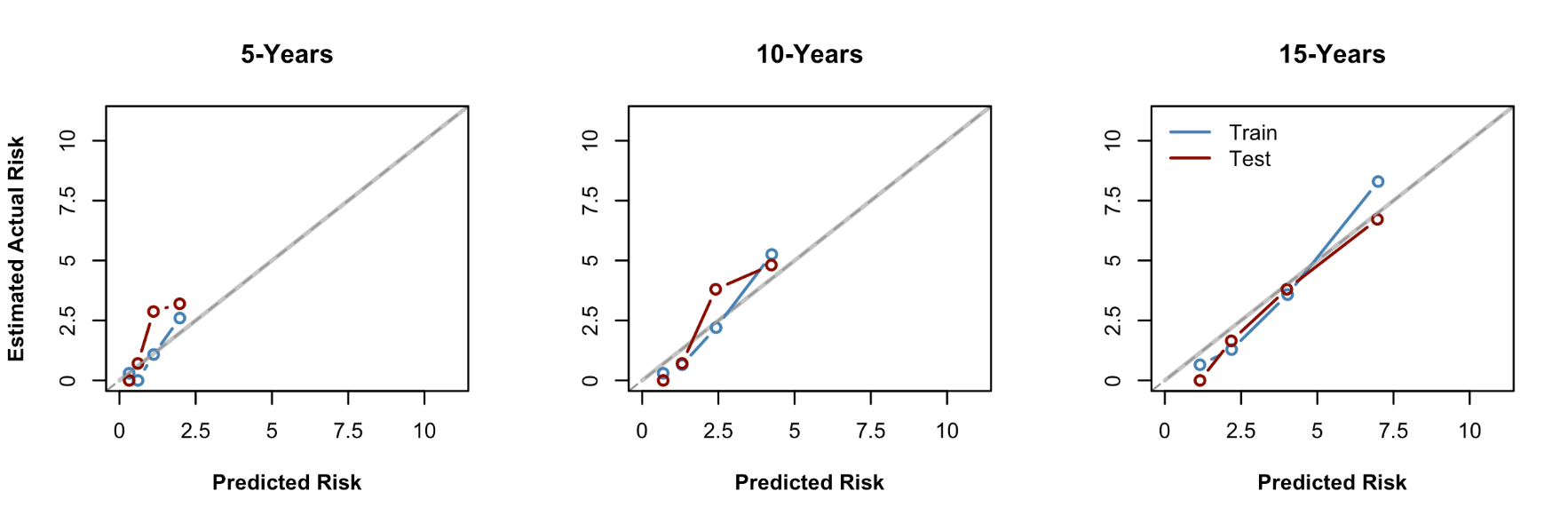

**Breast-Ovary**

**Breast-Pancreas**

**Prostate-Pancreas**

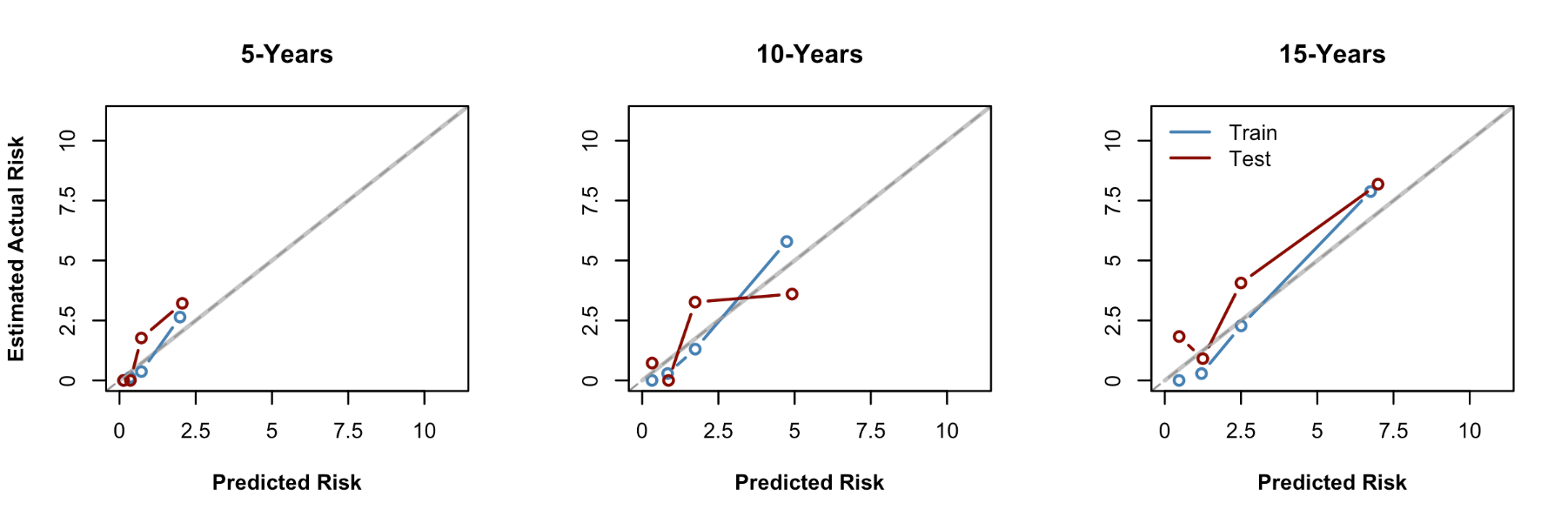

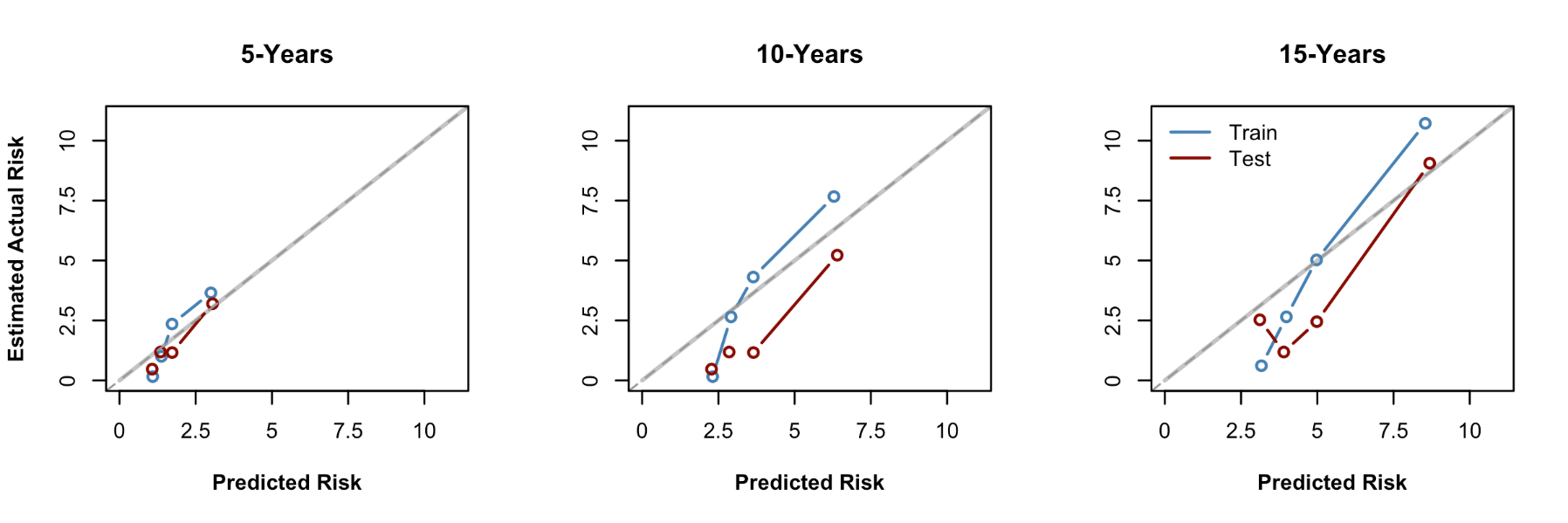

**Supplementary Figure 5. Calibration of predicted risk for second primary cancers across MPC-PREDICT models and time horizons**. Calibration plots are shown at 5, 10, and 15 years from the 1-year landmark date (1 year after first cancer diagnosis). Points represent actual risk within quartiles of predicted risk (each circle corresponding to increasing quartiles), plotted at the mean predicted risk within each quartile, with lines connecting quartiles for the training (blue) and validation (red) sets. The diagonal line indicates perfect calibration, where predicted and estimated actual risks are equal. Points above the diagonal indicate underprediction (actual risk exceeds predicted risk), whereas points below the diagonal indicate overprediction. In the validation set, O/E ratios for breast–ovary were 1.76, 1.17, and 0.94 at 5, 10, and 15 years; for breast–pancreas 1.59, 0.99, and 1.48; and for prostate–pancreas 0.85, 0.58, and 0.85. O/E variability in the prostate–pancreas model likely reflects the small validation sample size. Closest agreement was observed for the breast–ovary model, where calibration improved progressively over time.

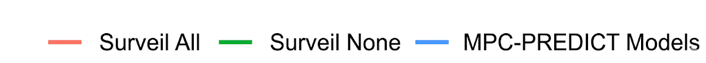

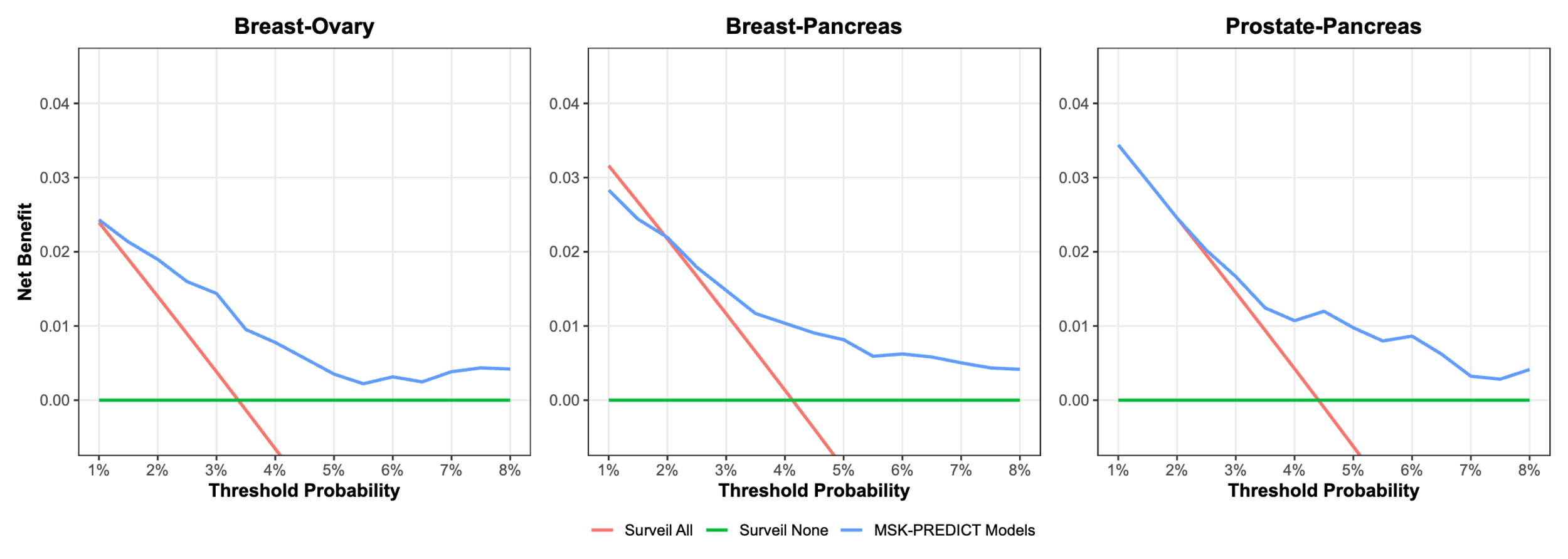

**Supplementary Figure 6.** **Decision curve analysis at 15 years for MPC-PREDICT models across cancer pairs**. Panels show net benefit for breast–ovary (A), breast–pancreas (B), and prostate–pancreas (C) cancer pairs. The MPC-PREDICT model (blue) is compared with *surveil-all* (red) and *surveil-none* (green) strategies. The y-axis represents net benefit, which incorporates the balance between true positives and false positives, weighted by the relative harm of unnecessary intervention. The x-axis shows the threshold probability, reflecting the risk at which a patient would be considered for surveillance or intervention. For example, a threshold of 5% corresponds to accepting approximately 1 true case per 20 individuals identified as high risk. Across all cancer pairs, the MPC-PREDICT models demonstrate higher net benefit than both *surveil-all* and *surveil-none* strategies across clinically relevant thresholds (approximately 3–8%), indicating potential clinical utility for risk-adapted surveillance. While the absolute gains in net benefit are modest, consistent with the low incidence of these specific second cancers, the models provide incremental improvement in identifying high-risk individuals while reducing unnecessary surveillance.

A

B

C

**Supplementary Figure 7.** **Insufficient performance of pathogenic variant status alone as a predictor for second cancer risk**. Time-dependent AUC at 5, 10, and 15 years from the 1-year landmark date (1 year after first cancer diagnosis; for colorectal–uterus, landmark was defined at colorectal diagnosis date due to low sample size) for Fine–Gray models using solely PV carrier status as predictors for the breast–ovary, breast–pancreas, prostate–pancreas, and colorectal–uterus cancer pairs. Points represent AUC estimates and error bars indicate 95% confidence intervals. Across all cancer pairs and time horizons, discrimination was mostly poor, with AUCs consistently in the range of approximately 0.53–0.63, highlighting the limited predictive utility of germline pathogenic variants alone.

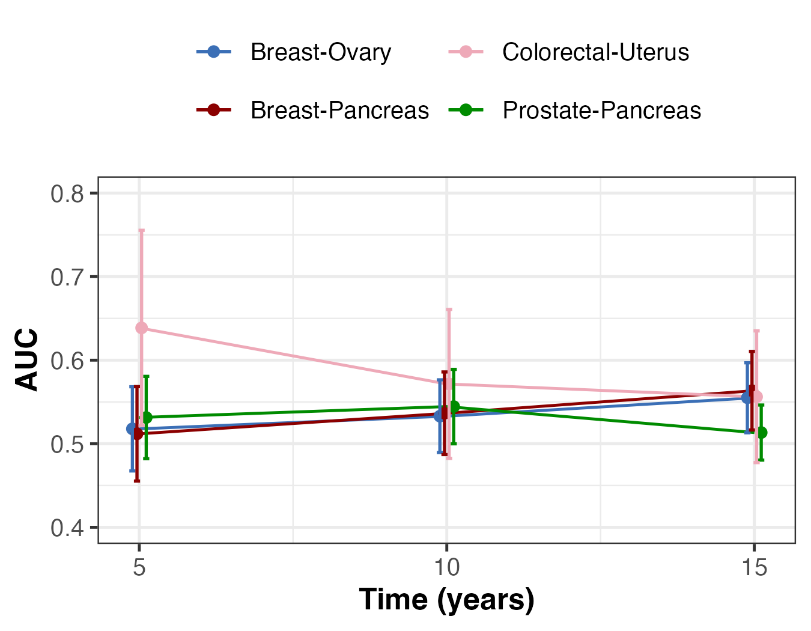

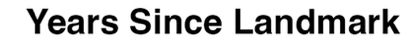
